# Optimal Prevalence Threshold For Guiding The Implementation Of Preventive Chemotherapy In Countries Endemic For Schistosomiasis: Synthesis Of Evidence From Mass Drug Administration Programmes For Developing This Tool

**DOI:** 10.1101/2021.05.10.21256643

**Authors:** Anthony Danso-Appiah, Paolo Eusebi, Nathan C. Lo, Massimiliano Orso, Kwadwo Owusu Akuffo, Fiona M. Fleming, Guo Jiangang, Pauline Mwinzi, Daniel G. Colley, Paul Hagan, M. Hassan Murad, Amadou Djirmay Garba

## Abstract

**Background:** The WHO-recommended prevalence thresholds for deciding schistosomiasis mass drug administration (MDA) are based on anecdotal evidence and may mislead. This study systematically synthesized evidence to generate a single optimal ‘global’ prevalence threshold that should guide schistosomiasis MDA programmes worldwide.

**Methods:** We searched several databases from 1978 to 31^st^ October 2021 without language restriction. Two reviewers selected studies, extracted data, and assessed risk of bias using relevant risk of bias tools, and resolved disagreements through discussion. The review followed best practices protocols and guidelines. Data were analysed and presented as prevalence reduction (PR) and relative risk (RR) for dichotomous outcomes or mean difference for continuous outcomes, each presented with their 95% confidence intervals (CI). Meta-regression of observations on prevalence rates and intensity of infection were performed to assess the effect of repeat MDA over time. Sensitivity analysis was performed to test the robustness of the results to the risk of bias components. The overall level of evidence was graded using GRADE.

**Findings:** Of the 1,232 studies retrieved, 38 met our inclusion criteria and 34 were included in the meta-analysis. No direct relation was observed between prevalence and intensity of infection. Praziquantel reduced prevalence of *S. mansoni* in school age children (SAC) at 12 months (RR 0.56, 95% CI 0.46 to 0.69; 14 studies, n=86,073); 24 months (RR 0.46; 95% CI 0.32 to 0.66; 14 studies; n=83,721); 36 months (RR 0.44, 95% CI 0.33 to 0.58; 7 studies, n=70,933) and 48 months (RR 0.25, 95% CI 0.11 to 0.59; 5 studies; n=27,483). Similarly for *S. haematobium,* there were reductions in prevalence in school age children (SAC) at 12 months (RR 0.38, 95% CI 0.28 to 0.52; 8 studies, n=37,868); at 24 months (RR 0.30; 95% CI 0.30─0.52; 7 studies; n=37,107); and 36 months (RR 0.39, 95% CI 0.21 to 0.71; 5 studies, n=28,146). There was no significant reduction in prevalence at 48 months (2 studies, n=10,954). Further analyses were performed from a series of prevalence thresholds created from the data at 5%, 10%, 15%, 20%, 30% and ≥40% and the results showed differences in the effect of MDA when each threshold was applied in the regression model. For annual MDA involving SAC, school-based treatment (SBT) appeared to perform better than community-wide treatment (CWT) in terms of prevalence reduction; but this could be subject to the frequency of treatment and retreatment applied in SAC compared to CWT. Using the optimal prevalence threshold of 10%, the model suggested it will take over 10 years to bring the prevalence of schistosomiasis to 1% for S. haematobium and up to 15 years for S. mansoni with repeated annual MDA.

**Interpretation:** This systematic review and meta-analysis provides evidence that 10% prevalence is the optimum that should be used as the **‘**standard global threshold’ for implementing MDA in endemic countries.

**Funding:** This work was commissioned and supported by the World Health Organization, Geneva, Switzerland as part of evidence-based schistosomiasis guideline development.

**Research in context:** *Evidence before this study:* Currently, the prevalence thresholds used in implementing mass drug administration within the preventive chemotherapy strategy for schistosomiasis control are based on anecdotal evidence and unreliable. We identified relevant studies regardless of language or publication status (published, unpublished, in press, and ongoing). We searched PubMed, CINAHL and LILACS from 1978 to 31^st^ October 2021 without language restriction. We also searched the Cochrane Infectious Diseases Group Specialized Register, CENTRAL (The Cochrane Library 2021), mRCT, Hinari, the WHO Library Database, Africa Journals Online and Google Scholar. Experts in the field of schistosomiasis were contacted, preprint repositories were searched and the reference lists of articles were reviewed for additional or unpublished data. This study was commissioned by the WHO to provide systematically synthesized evidence to inform on a single global prevalence threshold that should be applied by endemic countries when deciding MDA campaigns for the prevention and control of schistosomiasis.

*Added value of this study:* This is the first systematic review and meta-analysis commissioned by the WHO to determine a single prevalence threshold that should be employed by endemic countries for the implementation of global schistosomiasis mass drug administration. This study pooled data involving thousands of participants across thousands of villages from all endemic settings, making it unique in terms of statistical power and generalizability of the main findings and conclusions. The study used PICOS (P-population, I-intervention, C-comparator, O-outcomes and S-study) to formulate an appropriate review question, clear objectives, stringent inclusion and exclusion criteria as well as rigorous quality assessment and data synthesis, following strictly best practices for preparing and reporting systematic reviews. The search has been very comprehensive including all relevant electronic databases and non-electronic sources, done in close collaboration with experienced information specialist. The review process ensured meticulous attention to details, making the necessary effort to minimize bias, carrying out aspects of the review independently by the reviewers and addressing disagreements through discussions between the reviewers. Given the geographical variations, and differences in the levels of baseline endemicities, diagnostic criteria, age groups treated and follow-up times across studies, this necessitated robust sub-group analyses to detect any sub-group effects. We ran meta-regression analyses to identify any potentially useful trends, and tested the robustness of effect estimates from sensitivity analyses. We have assembled world-class experts from diverse backgrounds and geographical locations, including epidemiologists, evidence synthesis specialists, economists, allied health professionals, statisticians, biomedical scientists, clinicians and non-medical experts to produce this innovative, demand-driven, policy- and context-relevant systematic review and meta-analysis that will help guide policy and practice in the global control of schistosomiasis.

*Implications of all the available evidence:* Our review provides evidence that 10% baseline prevalence is the minimum optimal threshold that should be used to decide the implementation of MDA programmes in schistosomiasis endemic countries. Praziquantel is effective in reducing the prevalence of schistosomiasis at 12 months, but incremental benefit of repeated annual treatment appears to be minimal after 12 months. Effectiveness depends on several factors, which are difficult to disentangle, however, the rate at which prevalence decreases does not appear to be influenced by baseline intensity of infection and treatment approach (whether whole community or school-based). From exploratory analysis, intensity of infection appears to be more stable than prevalence for assessing outcome of MDA. Therefore, further research is needed to determine an optimal intensity threshold and compare it with prevalence threshold. In terms of policy, the difficulty in achieving elimination with mass drug administration alone means that integration of non-pharmacological interventions such access to clean water, improved sanitation, hygiene education (WASH) and snail control to complement MDA if elimination is to be achieved. This systematic review was registered in PROSPERO ̶ CRD422020221548.

## BACKGROUND

The ultimate goal of schistosomiasis control is to prevent new infection or transmission by interrupting the parasite’s lifecycle. This goal has been difficult to achieve, therefore, WHO guidelines and strategies have changed several times over the years.^1–4^ Since 2001, preventive chemotherapy (PC) has been the main control strategy applied in most endemic countries.^4–7^ Within the PC concept, endemic countries are urged to embark on mass drug administration (MDA) with Praziquantel (PZQ) at a single 40 mg/kg oral dose.^4^ Operational guidelines based on the World Assembly Resolution WHA54.19 with emphasis on morbidity control, using defined thresholds of infection prevalence as criteria for classifying at-risk populations, and selecting appropriate intervals of MDA were developed.^4^ Schools-based MDA was identified as the most cost-effective strategy. Since then, millions of school-aged children living in endemic countries have received multiple treatment with PZQ. Governments of endemic countries have partnered international non-governmental organizations, significant among them being the Schistosomiasis Control Initiative (SCI), the Schistosomiasis Consortium for Operational Research and Evaluation (SCORE), both funded by the Bill and Melinda Gates Foundation, the United States Agency for International Development (USAID), the British Department for International Development (DFID) and the Global Network for Neglected Tropical Diseases, to deliver MDA.^8–12^

In 2006, the operational details of the PC strategy were revised to expand the target population for MDA to include adults and special risk groups, for example, occupationally exposed individuals in high-risk areas where prevalence of infection in school age children reaches 50%.^5^ In 2012, following the release of the WHO Schistosomiasis Progress Report that reviewed the global progress towards control and elimination from 2001 to 2011 and set the agenda to guide control from 2012 to 2020, new goals were set.^7^ The WHO urged member states to increase control effort towards elimination of the disease as a public health problem.^7^ At the same time, the NTD road map and the London Declaration of NTD encouraged partners to pledge commitment towards reaching up to 100% of school age children living in endemic regions by 2020 and to eliminate the disease in some regions.^7, 13^ PZQ donation was to exceed 250 million tablets per year, and Countries and Programme Managers were encouraged to deliver PZQ to all at-risk populations at frequencies determined by the prevalence of infection.

Given that the scaling up of MDA has significantly brought intensity of infection in most endemic settings down to levels such that routinely used diagnostic tests are no longer sensitive to detect the infection, there was the need for sensitive diagnostic criteria. Therefore, the WHO commissioned a systematic review and meta-analysis that assessed the comparative accuracy of Point-of-care Circulating Cathodic Antigen (POC-CCA) and existing tools.^14^ Showing to be sensitive than existing tools, the WHO endorsed POC-CCA for the detection, monitoring and evaluation of *S. mansoni* MDA programmes (WHO Strategic Meeting in Geneva, 2016. A later study by Barenbold et al. showed similar findings.^15^

The current thresholds used in deciding MDA programmes vary, and are based on anecdotal evidence. The guideline stipulates that if baseline prevalence in school age children is ≥ 50% by parasitological methods (for intestinal and urogenital schistosomiasis) or ≥ 30% by questionnaires for history of haematuria or ≥ 60% by CCA in *S. mansoni*, all school age children (enrolled and not enrolled) should be treated once a year. At the same time, adults from special groups, for example, whose occupation predispose them to the acquisition of infection, to entire communities living in endemic areas considered to be at risk, should be treated. Additional interventions that should be applied, if operationally feasible, include improved water and sanitation, hygiene education (WASH) and snail control. For baseline prevalence among school age children ≥ 10% but <50% by parasitology (for intestinal and urogenital schistosomiasis) or <30% by questionnaire for history of haematuria or ≥15% but <60% by CCA with *S. mansoni*, all school age children (enrolled and not enrolled) should be treated once every two years with at least 50% of this age group treated each year. Adults considered to be at risk should also be treated and provision of wholesome water, sanitation and hygiene education (WASH) and snail control should be combined with MDA. If prevalence is <10% by parasitology or <15% by CCA in *S. mansoni* endemic areas, all school-aged children should be treated twice during their primary school period and MDA should cover at least 33% of this age group each year. Water, sanitation and hygiene education (WASH) and snail control should complement the control effort.

Until now, the WHO recommendations guiding the threshold at which MDA should be implemented have been based on consensus rather than systematically synthesized evidence. This systematic review was commissioned by the WHO to determine an optimal global (single) prevalence threshold that should guide endemic countries when deciding the implementation of PC programmes.

## METHODS

This systematic review used a well-formulated review question, clear objectives, stringent inclusion and exclusion criteria, rigorous quality assessment and quantitative synthesis, applying best practices for preparing systematic reviews. The search was comprehensive and done in close collaboration with experienced medical librarians. The review followed meticulous attention details, making the necessary effort to minimize bias at all the levels of the review process, carrying out aspects of the review independently by the reviewers and addressing disagreements through discussions.

### Search strategy and selection criteria

We identified all relevant studies regardless of publication status (published, unpublished, in press, preprints and ongoing). We searched PubMed, CINAHL and LILACS from 1978 to 31^st^ October 2021 without language restriction. We also searched the Cochrane Infectious Diseases Group Specialized Register, CENTRAL (The Cochrane Library 2021), mRCT, Hinari, the WHO Library Database, Africa Journals Online and Google Scholar. Experts in the field of schistosomiasis were contacted for additional or unpublished data and the reference lists of articles were reviewed. The search strategy and search terms have been reported in Table S1 (supplementary). Two reviewers independently selected studies, extracted data and assessed quality of the included studies using relevant risk of bias tooks^16–20^ Any discrepancies were resolved through discussion between the reviewers.

### Data analysis

The R software and Review Manager 5.4 were used for the analyses. Prevalence and absolute rate of events were calculated from all studies and relative risks from comparative studies and presented as prevalence ratios with their 95% CIs. Random-effects model was used with the meta-command for pooling data on treatment effect between baseline and first observation after MDA. Generalized linear mixed models were used for the meta-analysis and meta-regression of observations on prevalence rates of schistosomiasis before and after MDA. Different communities within a study and repeated follow-ups were each considered as separate data points. The interdependence between observations (communities and repeated follow-ups) within the same study or arms was accounted for by using linear mixed modelling (LMM). To this purpose, the lme4 package was used. Data were extracted to get the maximum level of details. In case information was available, each data row was an area within a study with a specific treatment for age group. The model contains random intercepts (the study units clustered within studies) and a random slope in time. This means that the rate at which schistosome infections decline after treatment is different from study-unit to study-unit. If a study unit has a negative random effect, then it decreases more quickly than the average. Whenever appropriate, we optimized the random effects, and the fixed-effects coefficients in the iteratively reweighted least-squares step by optimizing the parameter estimation. Wald 95 % CIs were derived. A p-value ≤0.05 was considered significant in all the analyses.

We assessed heterogeneity by inspecting forest plots for overlapping CIs and outlying data, and where heterogeneity was suspected, we conducted quantitative assessment from the Cochran’s Q test and *I^2^* statistic, setting the Chi^2^ test at a more sensitive threshold (P-value < 0.1) to indicate statistically significant heterogeneity.^21, 22^ Where significant heterogeneity was detected from the *I^2^* statistic, we carried out subgroup analyses based on clinical and methodological differences between the included studies. Where data were sufficient, we performed sensitivity analyses on the quality dimensions from GRADE and risk of bias elements to assess the robustness of the review findings to these factors. We used GRADE^23^ to grade the evidence by looking at 5 main evidence-based domains that might decrease quality of evidence, 1) study limitations, 2) inconsistency of results, 3) indirectness of evidence, 4) imprecision, and 5) publication bias. We also assessed the three domains that might increase quality of evidence including, 1) large magnitude of effect, 2) plausible confounding, which would reduce a demonstrated effect, and 3) dose-response gradient. For further information, visit GRADE website (http://www.gradeworkinggroup.org/).

## RESULTS

### Description of studies

The search retrieved 1,232 studies (1,186 studies from electronic databases and 46 studies from other sources including grey literature and contacts with experts), and 51 studies were excluded after deduplication. The remaining studies were screened and 1,094were excluded because they were not relevant to the topic. Full texts were obtained for 87 studies of which 49 studies were excluded because they were either non-MDA studies, reviews or did not provide treatment leaving with 38 studies that met our inclusion criteria. Four studies were excluded from the analysis because they presented only baseline data. Of the 34 studies included in the analysis, 15 assessed PZQ MDA on *S. mansoni*, six studies on *S. haematobium* and 13 studies assessed both *S. mansoni* and *S. haematobium*. No study conducted MDA on the other schistosome species (Fig. 1).

**Fig. 1.**
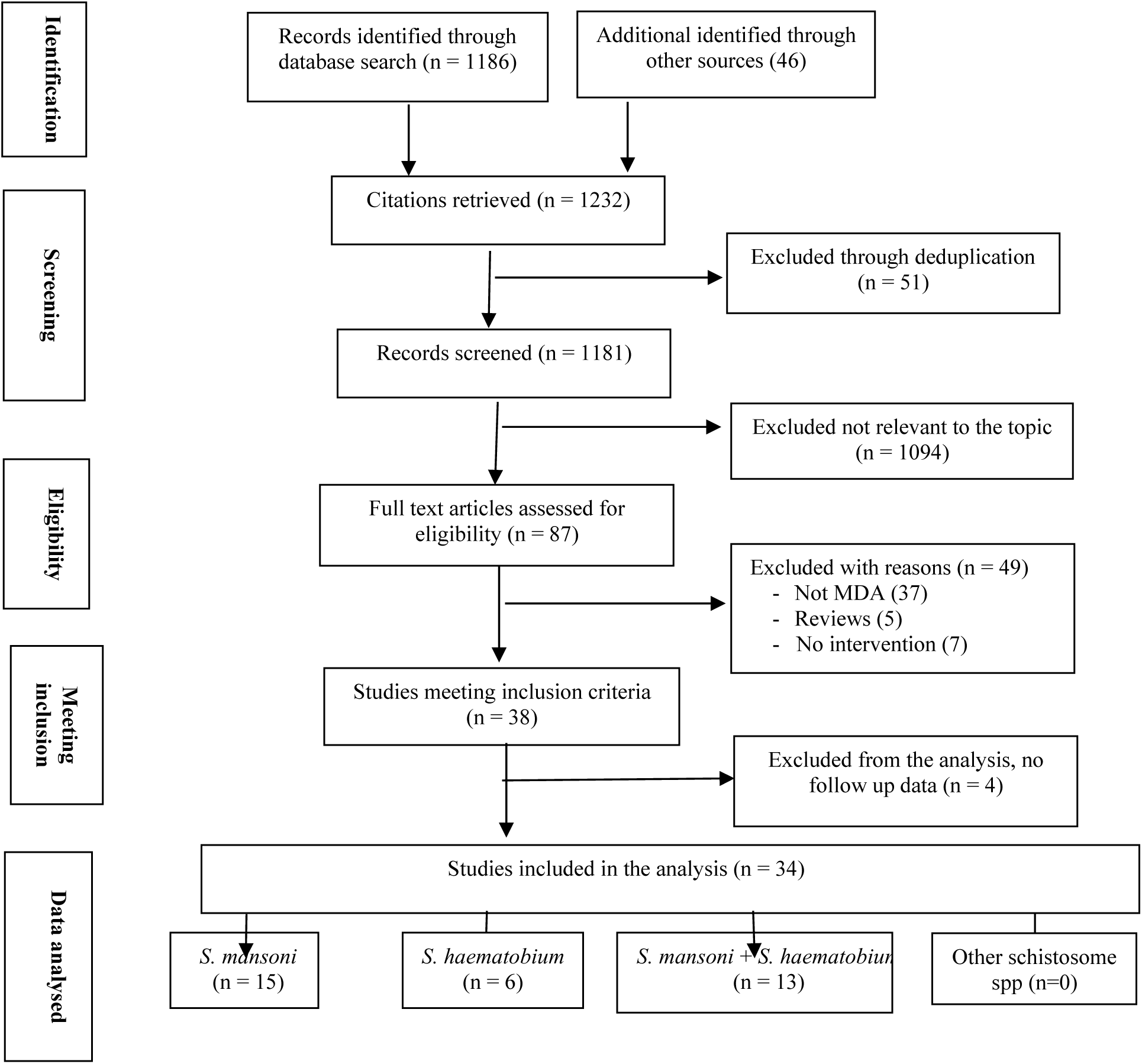
Flowchart of studies retrieved from relevant electronic databases and other sources (including contacts with experts in the field), number meeting the inclusion criteria and number included in the meta-analysis, by schistosome species.

### Description of the included studies

All the included MDA studies were conducted in Africa between 1988 and 2020 (see Characteristics of the included studies Table 1 and Table S2) Of the 34 studies included in the meta-analysis, 14 were conducted in West Africa ─ Burkina-Faso 2 studies,^24, 25^ Cote d’Ivoire 2 studies,^26, 27^ Mali 2 studies,^28, 29^ Niger 2 studies,^30, 31^ Senegal 2 studies,^32, 33^, Sierra Leone 2 studies,^34, 35^ Liberia 1 study,^36^ and Togo 1 study.^37^ Fourteen studies were conducted in East Africa, Kenya 8 studies,^38–45^ Tanzania 3 studies,^46–48^ Uganda 2 studies^49, 50^ and Ethiopia 1 study.^51^ Four studies were conducted in Central and Southern Africa ─ Burundi 1 study,^52^ Malawi 1 study,^53^ Mozambique 1 study,^54^ Rwanda 1 study,^55^ and the one study from North Africa was conducted in Sudan.^56^ Shen et al. conducted a two-country study in Kenya and Tanzania and reported combined data.^57^ We managed ten unpublished SCI-supported studies as independent MDA campaigns.^25, 29, 30, 36, 48, 50–53, 55^.

**Table 1.** Characteristics of the studies included in the meta-analysis

|  | Study ID | Country | Schisto species | Population | Diagnosis | Treatment | Follow-up (months) |
| --- | --- | --- | --- | --- | --- | --- | --- |
| 1 | Abudho et al., 2018<sup>38</sup> | Kenya | Sm | SAC | KK | PZQ+ALB | 48 |
| 2 | Assaré et al., 2016<sup>26</sup> | Cote d'Ivoire | Sm | SAC | KK | PZQ | 12 |
| 3 | Bah et al. 2019<sup>27</sup> | Cote d'Ivoire | Sm + Sh | SAC | KK, UF | PZQ | 36 |
| 4 | Brinkmann et al., 1988<sup>28</sup> | Mali | Sm + Sh | SAC | KK, UF | PZQ | 36 |
| 5 | Bronzan et al., 2018<sup>37</sup> | Togo | Sm + Sh | SAC | KK, UF | PZQ+ALB | 72 |
| 6 | Hamidou et al., 2018<sup>31</sup> | Niger | Sh | SAC | UF | PZQ | 60 |
| 7 | Hodges et al., 2012<sup>34</sup> | Sierra Leone | Sm | SAC | KK | PZQ+ALB | 6 |
| 8 | Karanja et al., 2017<sup>39</sup> | Kenya | Sm | SAC | KK | PZQ | 60 |
| 9 | Knopp et al., 2019<sup>46</sup> | Tanzania | Sh | SAC | UF | PZQ | 72 |
| 10 | Lee et al., 2015<sup>56</sup> | Sudan | Sm + Sh | SAC | KK, UF | PZQ | 6-9 |
| 11 | Moussa et al., 2016<sup>32</sup> | Senegal | Sm + Sh | SAC | KK, UF | PZQ | 6 |
| 12 | Mwandawiro et al., 2019<sup>40</sup> | Kenya | Sm + Sh | SAC | KK, UF | PZQ | 60 |
| 13 | Mwinzi et al., 2015<sup>41</sup> | Kenya | Sm | SAC | KK | PZQ+ALB | 6 |
| 14 | Njenga et al., 2014<sup>42</sup> | Kenya | Sh | SAC | UF | PZQ+ALB | 36 |
| 15 | Olsen et al., 2018<sup>47</sup> | Tanzania | Sm | SAC | KK | PZQ | 60 |
| 16 | Onkanga et al., 2016<sup>43</sup> | Kenya | Sm | SAC + Adults | KK | PZQ | 24 |
| 17 | Ouedraogo et al., 2016<sup>24</sup> | Burkina-Faso | Sh | SAC | UF | PZQ+ALB | 108 |
| 18 | Phillips et al., 2017<sup>54</sup> | Mozambique | Sh | SAC | UF | PZQ | 48 |
| 19 | SCI-supported MDA 1<sup>25</sup> | Burkina Faso | Sm+Sh | SAC | KK, UF | PZQ | 48 |
| 20 | SCI-supported MDA 2<sup>52</sup> | Burundi | Sm | SAC + Adults | KK | PZQ | 24 |
| 21 | SCI-supported MDA 3<sup>51</sup> | Ethiopia | Sm+Sh | SAC | KK, UF | PZQ | 72 |
| 22 | SCI-supported MDA 4<sup>36</sup> | Liberia | Sm+Sh | SAC | KK, UF | PZQ | 72 |
| 23 | SCI-supported MDA 5<sup>53</sup> | Malawi | Sm+Sh | SAC | KK, UF | PZQ | 48 |
| 24 | SCI-supported MDA 6<sup>29</sup> | Mali | Sm+Sh | SAC | KK, UF | PZQ | 48 |
| 25 | SCI-supported MDA 7<sup>30</sup> | Niger | Sm+Sh | SAC + Adults | KK, UF | PZQ | 48 |
| 26 | SCI-supported MDA 8<sup>55</sup> | Rwanda | Sm | SAC + Adults | KK | PZQ | 48 |
| 27 | SCI-supported MDA 9<sup>48</sup> | Tanzania | Sm+Sh | SAC + Adults | KK, UF | PZQ | 48-60 |
| 28 | SCI-supported MDA 10<sup>50</sup> | Uganda | Sm | SAC + Adults | KK, CCA | PZQ | 48 |
| 29 | Secor et al., 2020<sup>44</sup> | Kenya | Sm | SAC | KK | PZQ | 48 |
| 30 | Senghor et al., 2016<sup>33</sup> | Senegal | Sh | SAC | UF | PZQ | 60 |
| 31 | Sesay et al., 2014<sup>35</sup> | Sierra Leone | Sm | SAC | KK | PZQ | 48 |
| 32 | Shen et al., 2019<sup>57</sup> | Kenya+Tanzania | Sm | SAC | KK | PZQ | 36 |
| 33 | Sircar et al., 2018<sup>45</sup> | Kenya | Sm | SAC | KK | PZQ+ALB | 60 |
| 34 | Zhang et al., 2007<sup>49</sup> | Uganda | Sm | SAC | KK | PZQ+ALB | 48 |

PZQ = Praziquantel; ALB = Albendazole; SAC = School Age Children; Sm = *S. mansoni*; Sh = *S. haematobium*; KK = Kato-Katz; UF = Urine filtration, CCA = Circulating cathodic Antigen. The SCI-supported MDA data came from huge country surveys involving hundreds of villages in 10 Sub-Saharan African countries (Burkina Faso, Burundi, Ethiopia, Liberia, Malawi, Mali, Niger, Rwanda, Tanzania and Uganda). The SCI data were obtained from unpublished records.

The MDA programmes carried out so far involved *S. mansoni* and *S. haematobium*. Of the 34 studies included in the meta-analysis, 15 studies^26, 34, 35, 38, 39, 43–45, 47, 49, 52, 55, 57, 58^ were delivered in *S. manoni* endemic areas, six studies^24, 31, 33, 42, 46, 54^ in *S. haematobium* communities and 13 studies^25, 27–30, 32, 36, 37, 40, 48, 51, 53, 56^ were done in *S. mansoni* and *S. haematobium* mixed infection communities. All the *S. manoni* MDAs used the Kato-Katz’s (KK) stool technique for the detection of the infection,^26, 34, 35, 38, 39, 41, 43–45, 47, 49, 52, 55, 57^ whereas the *S. haematobium* MDs employed urine filtration (UF)^24, 31, 33, 42, 46, 54^ and for MDAs that involved mixed infections with S*. mansoni* and *S. haematobium*, all employed KK and UF for the detection of the infection.^25, 27–30, 32, 36, 37, 40, 48, 51, 53, 56^ Only one MDA study^50^ in *S. mansoni* endemic area in Uganda used combined KK and CCA test.

Twenty-eight studies^24–29, 31–40, 42, 44–47, 49, 51, 53, 54, 56–58^ involved SAC and six^30, 43, 48, 50, 52, 55^ involved SAC and adults. No MDA has been delivered to adults only. Twenty-six studies assessed PZQ alone^25–33, 35, 36, 39, 40, 43–48, 50–54, 56, 57^ and eight assessed PZQ combined with Albendazole.^24, 34, 37, 38, 41, 42, 45, 49^ Follow-up time after MDA ranged from six months to nine years (Table 1). Four studies^32, 34, 56, 58^ had follow-up time of 6-9 months, one study^26^ had 12 months follow-up, two studies^43, 52^ had 24 months follow up, four studies^27, 28, 42, 57^ had 36 months follow-up, eleven studies^25, 29, 30, 35, 38, 44, 49, 50, 53–55^ had 48 months, ten studies^31, 33, 36, 37, 39, 40, 45–47, 51^ had ^60–72^ months follow-up and one study^24^ had 108 months of follow-up. Mode of delivery of MDA differed between studies: majority of studies ^38,27,28,37,31,39,46,40,42,47,43,24,54,25,52,51,36,53,29,30,55,48,50,49,44,33,35,57,45^ delivered annual MDA for at least two years and two studies ^24, 31, 32, 39^ delivered bi-annual MDA. No study delivered biennial MDA. The majority of data came from the SCI and SCORE–supported MDA campaign programmes. SCI provided huge unpublished data,^25, 29, 30, 36, 48, 50–53, 55^ and we made sure to avoid duplication of data that have been published from the same data. The characteristics of the studies included in the review have been described in Table S2(supplementary).

### Prevalence versus intensity of infection

We estimated the relationship between prevalence and intensity of infection in both *S. mansoni* and *S. haematobium* across different endemicities and settings and the results showed inverse quadratic relationships for both *S. mansoni* and *S. haematobium*, with *S. mansoni* showing slightly steeper curve than *S. haematobium* (Fig. 2 A and B). At 10 epg intensity, the corresponding prevalence ranged from 1% to about 35%, at 100 epg, prevalence ranged from 1% to 80%, at 1,000 epg, prevalence ranged from 1% to about 98% and for epg 100-1,000, prevalence ranged from 1% to 100% for *S. mansoni* (Fig. 2A). Clearly, there is no direct correlation between intensity and prevalence of infection. The same was demonstrable for *S. haematobium,* 10 eggs per 10mL corresponded with prevalence ranging from about 5% to 75%, 100 eggs per 10mL corresponded with about 5% to 85% prevalence and 100-1,000 eggs per 10mL corresponded with 5% to 95% prevalence (Fig. 2B). There is no direct proportionality between intensity and prevalence of infection as demonstrated for *S. mansoni* and *S. haematobium* infections from this huge data obtained from millions of participants in MDA campaigns.

**Fig. 2.**
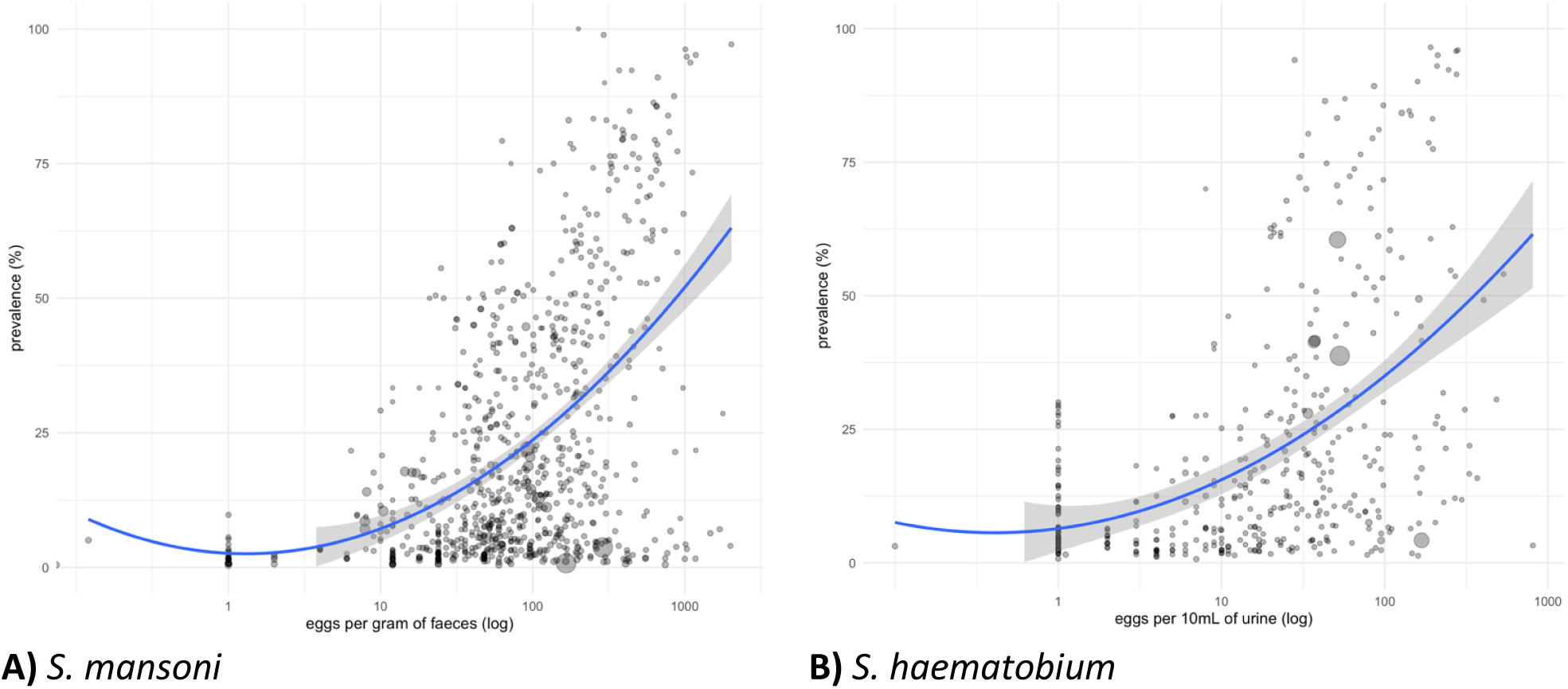
Relationship between prevalence (as proportion) and infection intensity of infection expressed as EPG (eggs per gram of faeces) for *S. mansoni* (**A**) or eggs per 10mL of urine for *S. haematobium* (**B**). The data points (black circles) came from 28 large studies involving thousands of villages and millions of inhabitants who received PZQ for *S. mansoni* and 22 studies of thousands of villages and millions of inhabitants treated with PZQ for *S. haematobium*. Given that egg output does not follow a normal distribution, EPG and eggs/10mL values (x-axis) have been reported on a log scale. A quadratic polynomial regression was fitted to depict the relationship between prevalence and intensity of infection, with their 95% CIs (grey area). The size of the circles is proportional to the size of the population.

### Meta-analysis of the effect of MDA

We assessed the effect of annual PZQ MDA on prevalence of *S. mansoni* in school children (SAC) in a meta-analysis between baseline and different follow-up times at 12, 24, 36 and 48 months (Fig. 3). Fourteen studies^25, 26, 29, 30, 36, 38, 43, 48–53, 55^ investigated SAC annual MDA through school-based strategy, and the results showed a reduction in prevalence at 12 months (RR 0.56, 95% CI 0.46 to 0.69; 14 studies, 45,510 school children, *I^2^*=97%); at 24 months (RR 0.46; 95% CI 0.32 to 0.66; 14 studies; 41,138 school children, *I^2^*=99%); at 36 months (RR 0.44, 95% CI 0.33 to 0.58; 7 studies, 40,698 school children, *I^2^*=92%) and at 48 months (RR 0.25, 95% CI 0.11 to 0.59; 5 studies; 17,745 school children, *I^2^*=99%), translating into 44%, 54%, 56% and 75% prevalence reductions, respectively

**Fig. 3.**
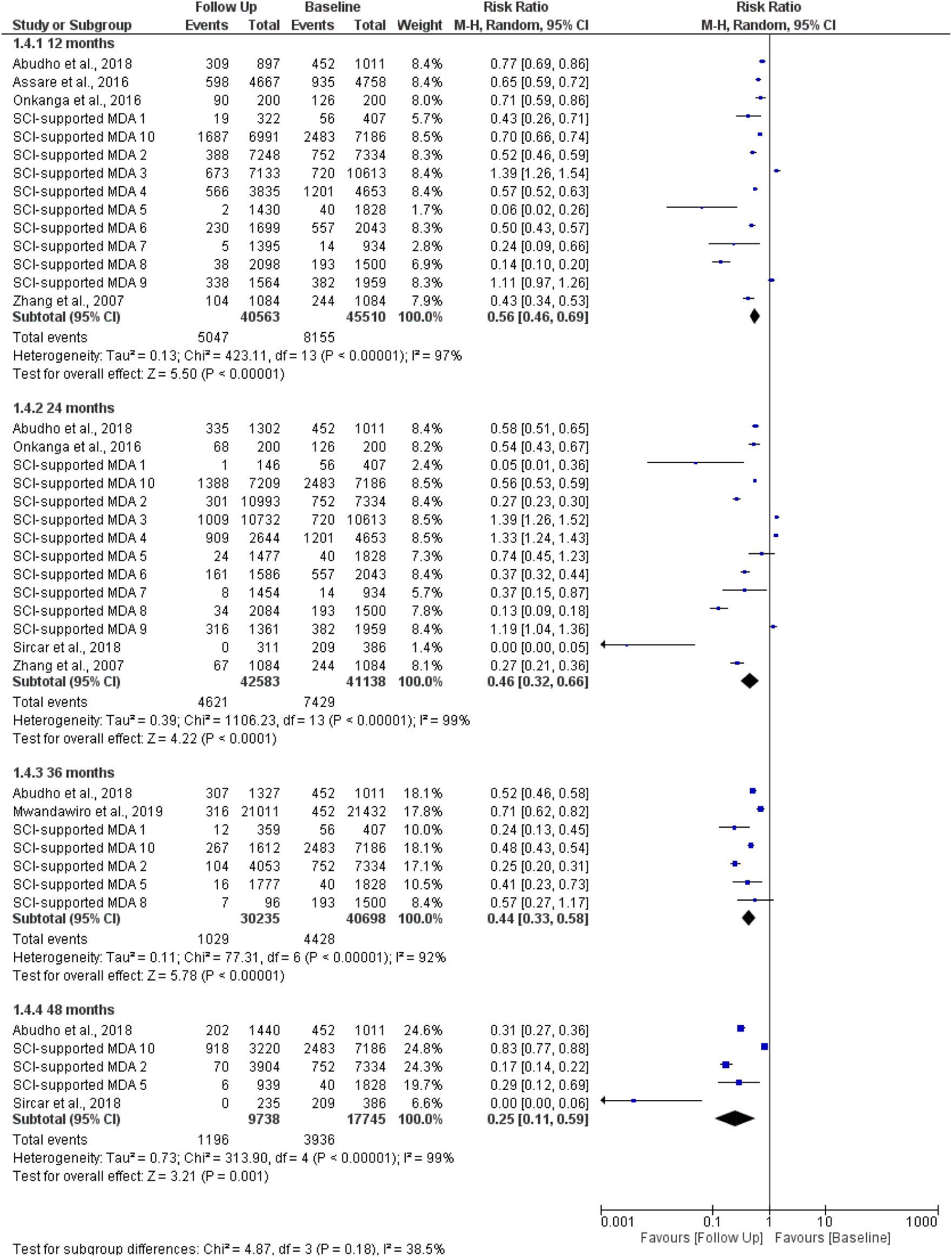
Forest plot of effectiveness of PZQ 40 mg/kg single dose in SAC for *S. mansoni*. The vertical line denotes the point of no difference between baseline and follow up prevalence, therefore, if the 95% CI includes the value 1, it means there is no statistically significant difference between baseline and follow up prevalence after MDA. Each study has a square denoting the ‘effect estimate’ for that particular study with the horizontal line in the middle of the square denoting the 95% CI. The size of the square is proportional to the size of the study and the bigger the study the narrower the CI. Each study is assigned a weight based on contribution to the pooled estimate (diamond shape).

In this meta-analysis, we assessed the effect of annual PZQ MDA in SAC between baseline and successive follow-up times for *S. haematobium* infection at 12, 24, 36 and 48 months (Fig. 4). Eight studies^25, 29, 30, 36, 48, 51, 53, 54^ investigated annual SAC MDA, and the results showed a statistically significant reduction in the prevalence of infection at 12 months (RR 0.38, 95% CI 0.28 to 0.52; 8 studies, 20,040 school children, *I^2^*=99%); at 24 months (RR 0.39; 95% CI 0.30 to 0.52; 8 studies; 20,040 school children, *I^2^*=98%); at 36 months (RR 0.39, 95% CI 0.21 to 0.71; 5 studies, 13,689 school children, *I^2^*=99%) and although not statistically significant at 48 months, PZQ MDA led to an overall reduction in prevalence (RR 0.38, 95% CI 0.14 to 1.07; 2 studies; 6,274 school children, *I^2^*=98%), translating into 62%, 61%, 61% and 62% prevalence reductions, respectively.

**Fig. 4.**
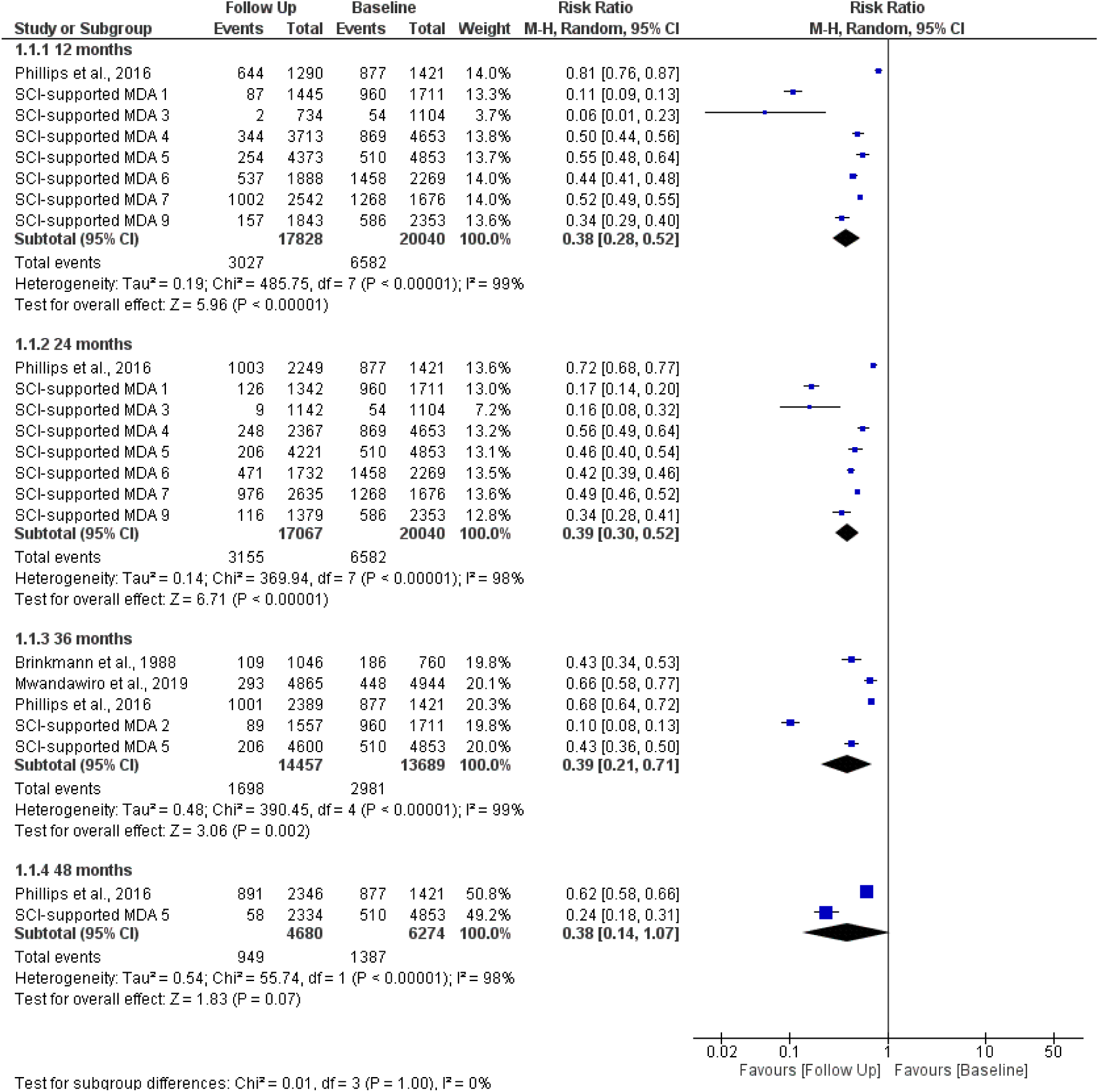
Forest plot of the effectiveness of MDA with PZQ 40 mg/kg single dose in SAC for *S. haematobium*. The vertical line denotes the point of no difference between baseline and follow up prevalence. For further explanation of the forest plot see Fig. 3.

Meta-analysis of effect of school-based treatment (SBT) in adults living in *S. mansoni* endemic settings at baseline and successive follow-up times at 12, 24, 36 and 48 months (Fig. 5). Four studies^48, 50, 52, 55^ that investigated this showed reductions in prevalence at 12 months (RR 0.55, 95% CI 0.38 to 0.82; 4 studies, 2,344 participants, *I^2^*=88%); at 24 months (RR 0.25; 95% CI 0.14 to 0.46; 4 studies; 2,344 participants, *I^2^*=89%); at 36 months (RR 0.31, 95% CI 0.10 to 0.93; 3 studies, 1,356 participants, *I^2^*=74%) and at 48 months (RR 0.37, 95% CI 0.19 to 0.70; 2 studies; 1,16944 participants, *I^2^*=19%).

**Fig. 5.**
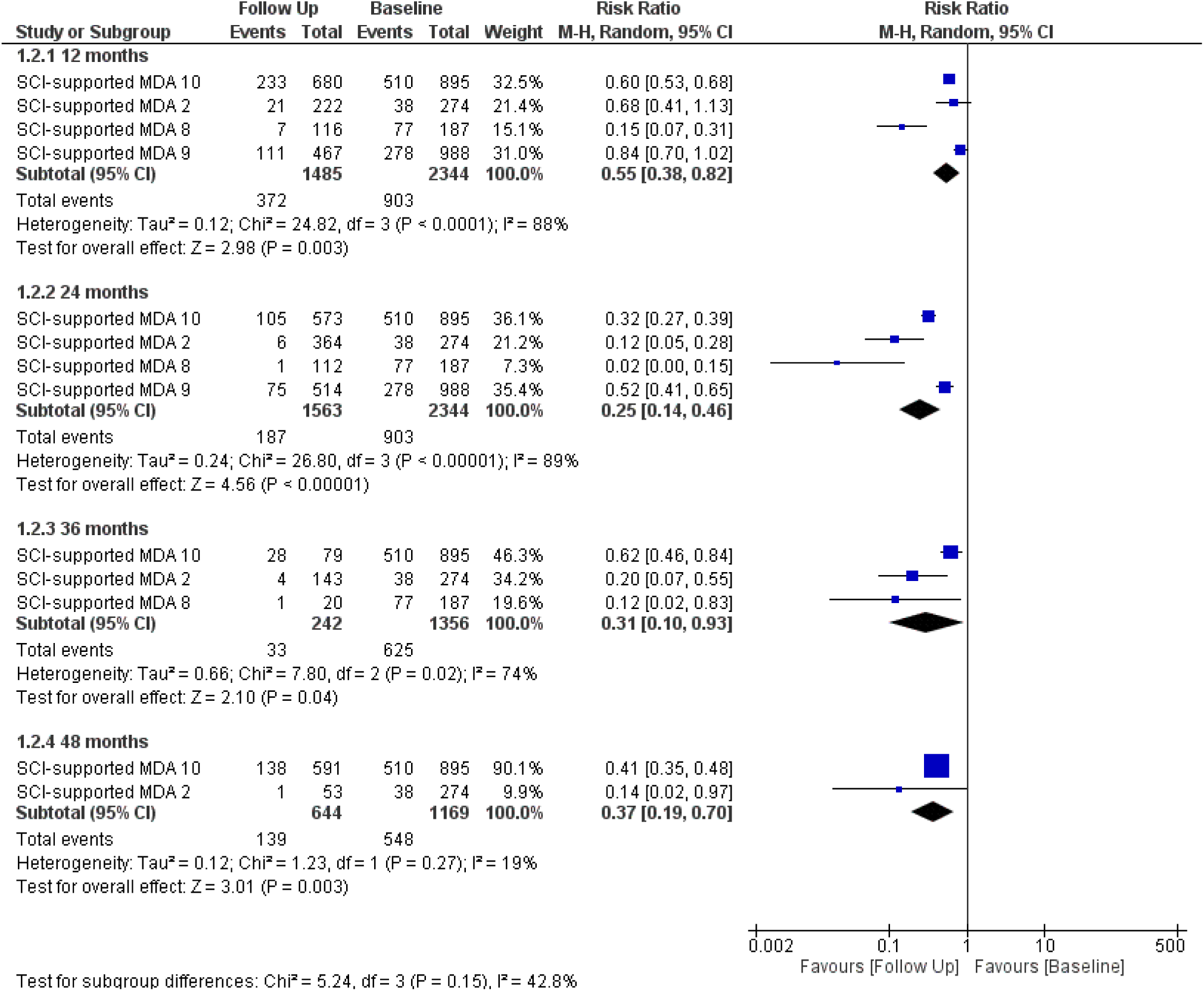
Effectiveness of MDA with PZQ 40 mg/kg single dose in adults with *S. mansoni* infection at baseline and successive follow up times. For further explanation of the forest plot, see Fig. 3.

The meta-analysis of the effect of annual Community-Wide Treatment (CWT) in adults that compared baseline and follow-up prevalence of infection after 12, 24, 36 and 48 months of has been presented (Fig. 6). Two studies^43, 49^ investigated CWT and showed reduction in prevalence of infection at various follow up time: at 12 months (RR 0.77, 95% CI 0.68 to 0.88; 2 studies, 820 participants, *I^2^*=52%); at 24 months (RR 0.63; 95% CI 0.41 to 0.99; 3 studies; 1,236 participants, *I^2^*=94%); at 36 months (RR 0.40, 95% CI 0.30 to 0.54; 2 studies, 2,108 participants, *I^2^*=85%) and at 48 months (RR 0.37, 95% CI 0.19 to 0.70; 2 studies; 1,169 participants, *I^2^*=19%), 23%, 37%, 60% and 63% prevalence reductions, respectively.

**Fig. 6.**
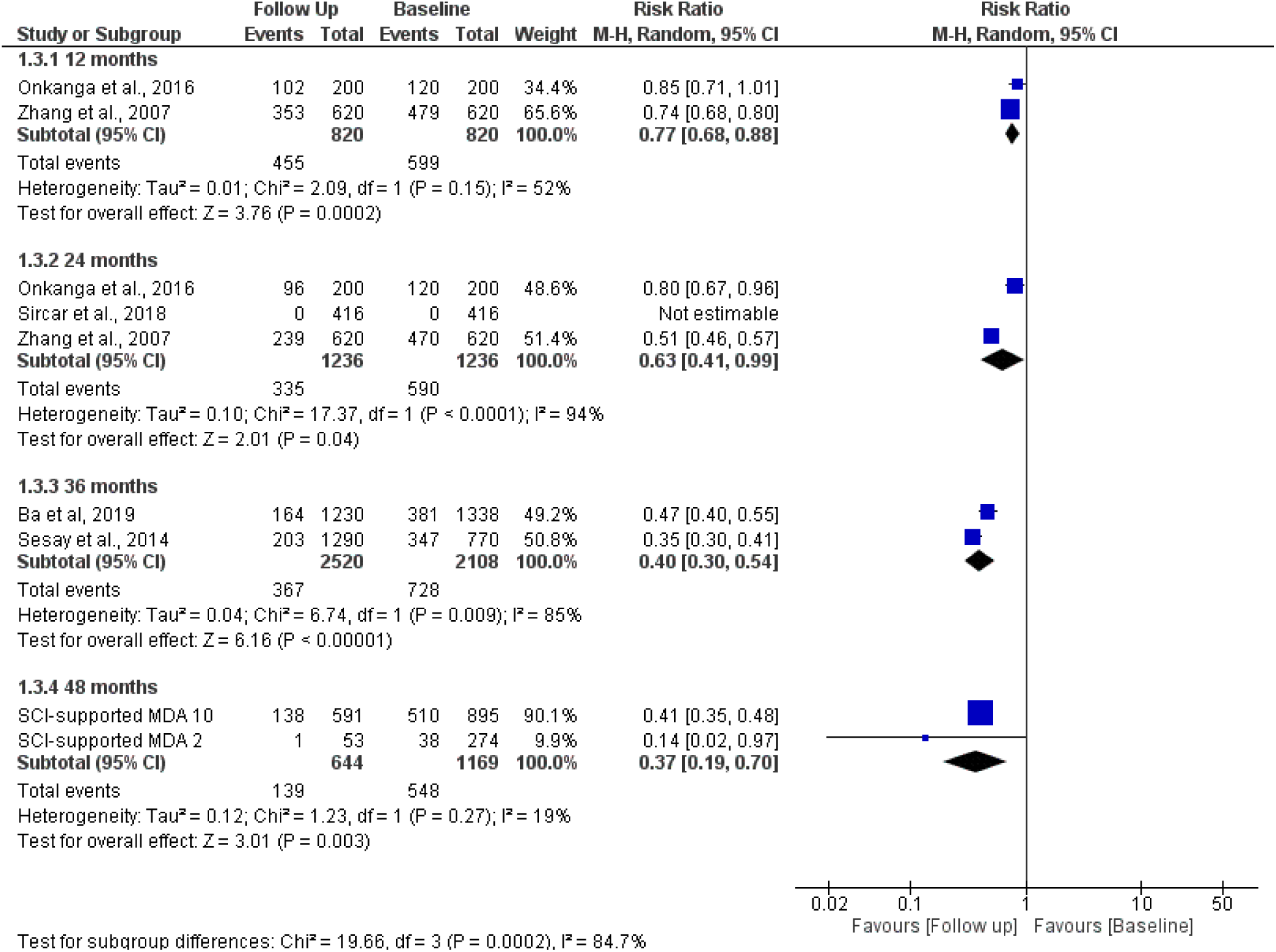
Forest plot of effect of single 40 mg/kg oral dose in adults with *S. mansoni* treated through community-side mass drug administration campaigns (see Fig. 3 for further explanation of the figure).

Praziquantel was effective in reducing prevalence of *S. mansoni* (PR=0.83, 95% CI = 0.82 to 0.83) and *S. haematobium* (PR=0.76, 95% CI = 0.75 to 0.76), translating into a 17% annual reduction for *S. mansoni* and 24% annual reduction for *S. haematobium* in MDA programmes in a very highly powered meta-analysis, deriving the estimates from generalized linear model (GLM) for longitudinal data (Table S3). There was a sharp drop of prevalence after MDA but the decrease became less prominent with the curves appearing to be leveling off over time, particularly for *S. mansoni*. Further analysis showed that annual MDA for SAC and adults for *S. mansoni* and *S. haematobium* led to a decreased in prevalence rate (Fig. 7), more prominently in SAC (A and C) than adults (B and D). Prevalence after MDA in adults showed sharp decline of up to 2 years and then started to increase (B) whereas for *S. haematobium* after initial drop, prevalence appears to level off (D).

**Fig. 7.**
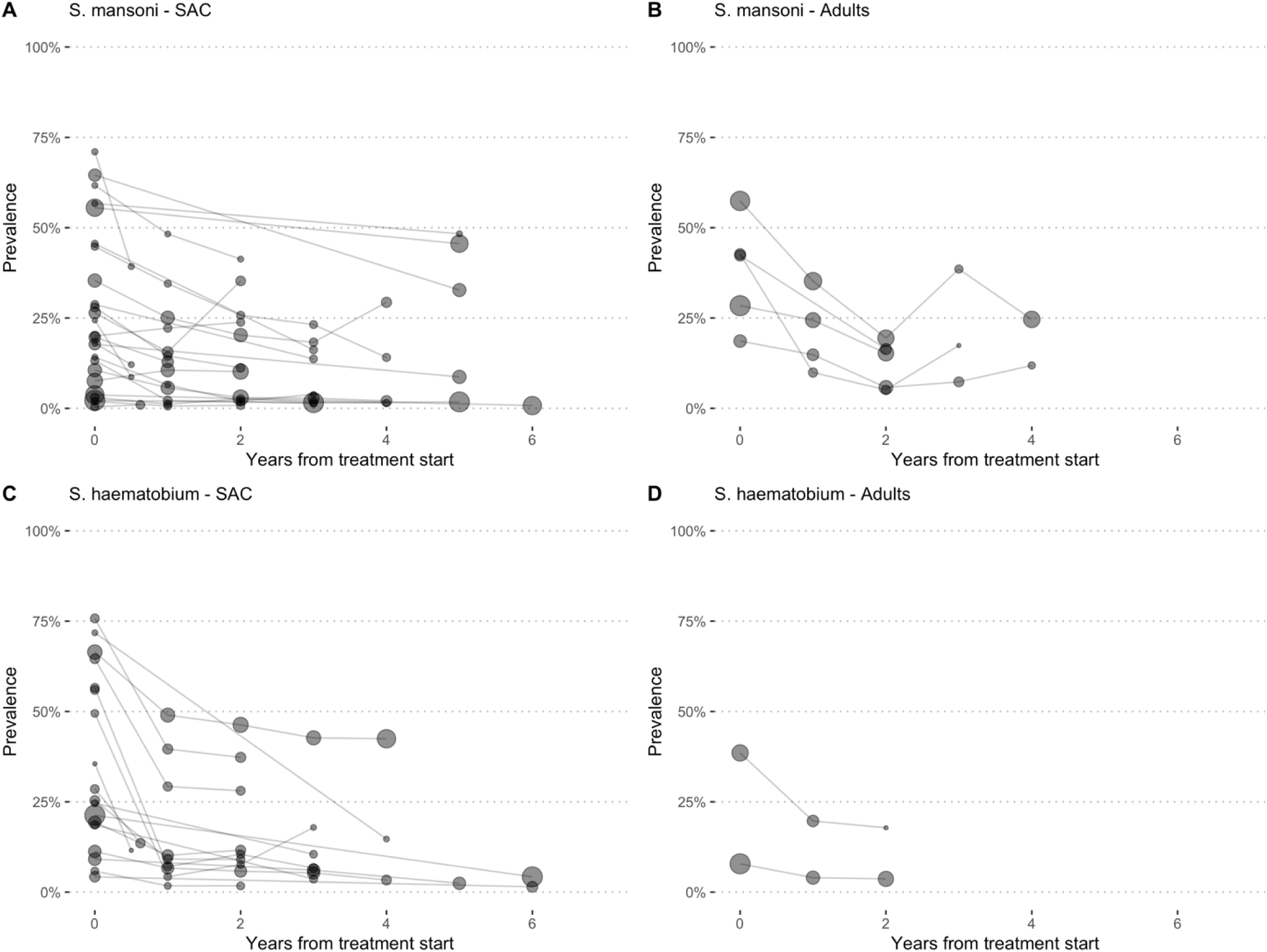
Prevalence Rates over time points following MDA of adults are plotted for studies that investigated *S. mansoni* in SAC (A), *S. mansoni* in adults (B), *S. haematobium* SAC (C) and *S. haematobium* adults (D). The size of the circles is proportional to the size of the population at each follow-up of assessment. Follow up was up to six years.

The meta-regression on prevalence ratio to assess effectiveness of the two main MDA strategies — community-wide treatment (CWT) and school-based treatment (SBT) according to prevalence thresholds showed that CWT did not demonstrate any obvious differences in prevalence reductions estimates for *S. mansoni* for school children at 5% threshold (PR=0.852, 95% CI 0.852 to 0.871, 5 studies, 9,104 school children), at 10% threshold (PR=0.852, 95% CI 0.833 to 0.8871, five studies, 9,104 school children) and at 15% threshold (PR=0.852, 95% CI 0.852 to 0.871, five studies, 9,104 school children); 14.8%, 14.8% and 14.8% prevalence reductions, respectively.

School-based treatment also did not produce different prevalence reduction at 5% threshold (PR=0.783, 95% CI 0.775 to 0.79, twenty studies, 181 arms, 123,045 school children), at 10% threshold (PR=0.778, 95% CI 0.771 to 0.786, nineteen studies, 141 arms, 97,943 school children) and at 15% threshold (PR=0.769, 95% CI 0.762 to 0.777, eighteen studies, 84,647 school children); 21.7%, 22.2% and 23.1% prevalence reductions, respectively. The results did not differ for SBT for adults at 5%, 10% and 15% thresholds: (PR=0.665, 95% CI 0.63 to 0.702, four studies, 32 arms, 5,358 participants), (PR=0.661, 95% CI 0.626 to 0.698, four studies, 29 arms, 5,065 participants) and (PR=0.66, 95% CI 0.625 to 0.697, four studies, 26 arms, 4,879 participants); 30%, 30% and 30% prevalence reductions, respectively. Similar results were obtained for SBT or CWT for *S. haematobium* (Table 2).

**Table 2.** Meta-regression results of Prevalence Ratios (PRs) between baseline and follow-up after treatment, with their 95% CIs for various thresholds by MDA type

| Schisto. species | No. of people | No. of studies | Arms | Age group | MDA type | Threshold | PR | PR_95_CI |
| --- | --- | --- | --- | --- | --- | --- | --- | --- |
| *S. mansoni* | 9,104 | 5 | 5 | SAC | Annual CWT | 0.05 | 0.852 | 0.852 (0.833-0.871) |
| *S. mansoni* | 9,104 | 5 | 5 | SAC | Annual CWT | 0.1 | 0.852 | 0.852 (0.833-0.871) |
| *S. mansoni* | 9,104 | 5 | 5 | SAC | Annual CWT | 0.15 | 0.852 | 0.852 (0.833-0.871) |
| *S. mansoni* | 123,045 | 20 | 181 | SAC | Annual SBT | 0.05 | 0.783 | 0.783 (0.775-0.79) |
| *S. mansoni* | 97,943 | 19 | 141 | SAC | Annual SBT | 0.1 | 0.778 | 0.778 (0.771-0.786) |
| *S. mansoni* | 84,647 | 18 | 121 | SAC | Annual SBT | 0.15 | 0.769 | 0.769 (0.762-0.777) |
| *S. mansoni* | 5,358 | 4 | 32 | Adults | Annual SBT | 0.05 | 0.665 | 0.665 (0.63-0.702) |
| *S. mansoni* | 5,065 | 4 | 29 | Adults | Annual SBT | 0.1 | 0.661 | 0.661 (0.626-0.698) |
| *S. mansoni* | 4,879 | 4 | 26 | Adults | Annual SBT | 0.15 | 0.66 | 0.66 (0.625-0.697) |
| *S. haematobium* | 20,646 | 4 | 4 | SAC | Annual CWT | 0.05 | 0.821 | 0.821 (0.804-0.838) |
| *S. haematobium* | 20,646 | 4 | 4 | SAC | Annual CWT | 0.1 | 0.821 | 0.821 (0.804-0.838) |
| *S. haematobium* | 13,600 | 3 | 3 | SAC | Annual CWT | 0.15 | 0.71 | 0.71 (0.69-0.731) |
| *S. haematobium* | 106,912 | 13 | 123 | SAC | Annual SBT | 0.05 | 0.743 | 0.743 (0.736-0.751) |
| *S. haematobium* | 99,931 | 13 | 105 | SAC | Annual SBT | 0.1 | 0.743 | 0.743 (0.736-0.751) |
| *S. haematobium* | 91,555 | 13 | 91 | SAC | Annual SBT | 0.15 | 0.743 | 0.743 (0.735-0.75) |
| *S. haematobium* | 1,722 | 2 | 6 | Adults | Annual SBT | 0.05 | 0.575 | 0.575 (0.475-0.697) |
| *S. haematobium* | 1,722 | 2 | 6 | Adults | Annual SBT | 0.1 | 0.575 | 0.575 (0.475-0.697) |
| *S. haematobium* | 1,284 | 2 | 5 | Adults | Annual SBT | 0.15 | 0.569 | 0.569 (0.462-0.701) |

Arms = number of communities\_arms; PR = prevalence ratio; SAC = school age children; CWT = community wide treatment; SBT = school-based treatment; MDA = mass drug administration; PR = prevalence ratio; CI = confident interval; Estimates are derived from General Linear Model (GLM)

We also explored PR by type of MDA (SBT or CWT) and baseline prevalence (low, moderate and high) in *S. mansoni* and *S. haematobium* endemicity settings (Table 3). The results showed that in *S. mansoni* endemic settings, providing annual SBT for SAC, led to decreases in prevalence: low prevalence setting (PR=0.878, 95% CI 0.86 to 0.897, twelve studies), moderate prevalence (PR=0.706, 95% CI 0.691 to 0.721, fourteen studies) and high prevalence (PR=0.682, 95% CI 0.662 to 0.703, nine studies), translating to 10%, 27.8% and 30% for low, moderate and high baseline prevalence, respectively. Three studies that assessed annual CWT in settings with high baseline prevalence found pooled prevalence reduction of 44% (PR=0.507, 95% CI 0.457 to 0.562). One study investigated combined SBT-CWT in a moderate prevalence setting and showed a reduction of 25% whilst another combined CWT-SBT study in a high prevalence setting found a reduction of 43%. Annual SBT in low *S. haematobium* prevalence setting showed outcome similar to that of *S. mansoni* (PR=0.761, 95% 0.722 to 0.803, seven studies): moderate baseline prevalence (PR=0.721, 95% CI 0.712 to 0.731, nine studies) and high baseline prevalence (PR=0.774, 95% CI 0.762 to 0.787, seven studies). Two studies annual CWT in moderate prevalence setting and found better pooled reduction than children (PR=0.585, 95% CI 0.477 to 0.719). Annual CWT involving whole population in moderate and high *S. haematobium* prevalence settings showed higher reductions but these came from a single study each (Table 3).

**Table 3.** Meta-regression results in terms of Prevalence Ratios with 95% CIs per year of treatment and baseline prevalence or type of treatment

| Schisto species | Age | MDA type | Baseline Prevalence | PR (95% CI) | Studies |
| --- | --- | --- | --- | --- | --- |
| *S. mansoni* |  |  |  |  |  |
|  | SAC | Annual SBT | Low | 0.878 (0.86 to 0.897) | 12 |
|  | SAC | Annual SBT | Moderate | 0.706 (0.691 to 0.721) | 14 |
|  | SAC | Annual SBT | High | 0.682 (0.662 to 0.703) | 9 |
|  | SAC | Annual CWT | High | 0.507 (0.457 to 0.562) | 3 |
|  | SAC | Annual SBT-CWT | Moderate | 0.678 (0.614 to 0.75) | 1 |
|  | SAC | Annual SBT-CWT | High | 0.514 (0.463 to 0.57) | 1 |
|  | Adults | Annual SBT | Low | 0.936 (0.716-1.222) | 2 |
|  | Adults | Annual SBT | Moderate | 0.699 (0.637 to 0.768) | 4 |
|  | Adults | Annual SBT | High | 0.659 (0.615 to 0.707) | 3 |
| *S. haematobium* |  |  |  |  |  |
|  | SAC | Annual SBT | Low | 0.761 (0.722 to 0.803) | 7 |
|  | SAC | Annual SBT | Moderate | 0.721 (0.712 to 0.731) | 9 |
|  | SAC | Annual SBT | High | 0.774 (0.762 to 0.787) | 7 |
|  | Adults | Annual SBT | Moderate | 0.585 (0.477 to 0.719) | 2 |
|  | All | Annual CWT | Moderate | 0.471 (0.396 to 0.561) | 1 |
|  | All | Annual CWT | High | 0.562 (0.513 to 0.615) | 1 |
|  | SAC | Annual CWT | High | 0.828 (0.811 to 0.845) | 3 |

SAC = school age children; CWT = community wide treatment; SBT = school-based treatment; MDA = mass drug administration; PR = prevalence ratio; CI = confident interval; Estimates are derived from General Linear Model (GLM).

### Prevalence reduction following MDA

Clearly, PZQ was effective in reducing the prevalence of schistosomiasis by at least 25% per year for *S. mansoni* and 23% per year for *S. haematobium*. Effectiveness depends on several factors, which are difficult to disentangle but it is obvious that rate of prevalence reduction is influenced by baseline endemicity for SAC and adults in *S. mansoni* and *S. haematobium* endemic settings with areas categorized as high endemicity showing the highest drop followed by moderate endemicity (Fig. 8). In low endemicity areas, MDA led to marginal prevalence reduction over time. Nevertheless, rate of decrease slows down over time and prevalence for high, moderate and low endemicities for SAC and adults approximate each other in what appears to be a levelling off effect for both *S. mansoni* and *S. haematobium,* but this effect is not that apparent with SAC in *S. haematobium* settings (Fig. 8). For *S. mansoni*, repeated annual MDA brought infection levels down to about 1-2% after 10 years for three endemicity categories (low, moderate and high). Similar observation was seen with low and moderate endemcity of *S. hamatobium* but not high endemicity for SAC which reached around 10% prevalence after 10 years of repeated MDA. Similar results were observed regardless of the MDA strategy employed─ whether CWT, SBT or combined SBT-CWT (Fig. 9).

**Fig. 8.**
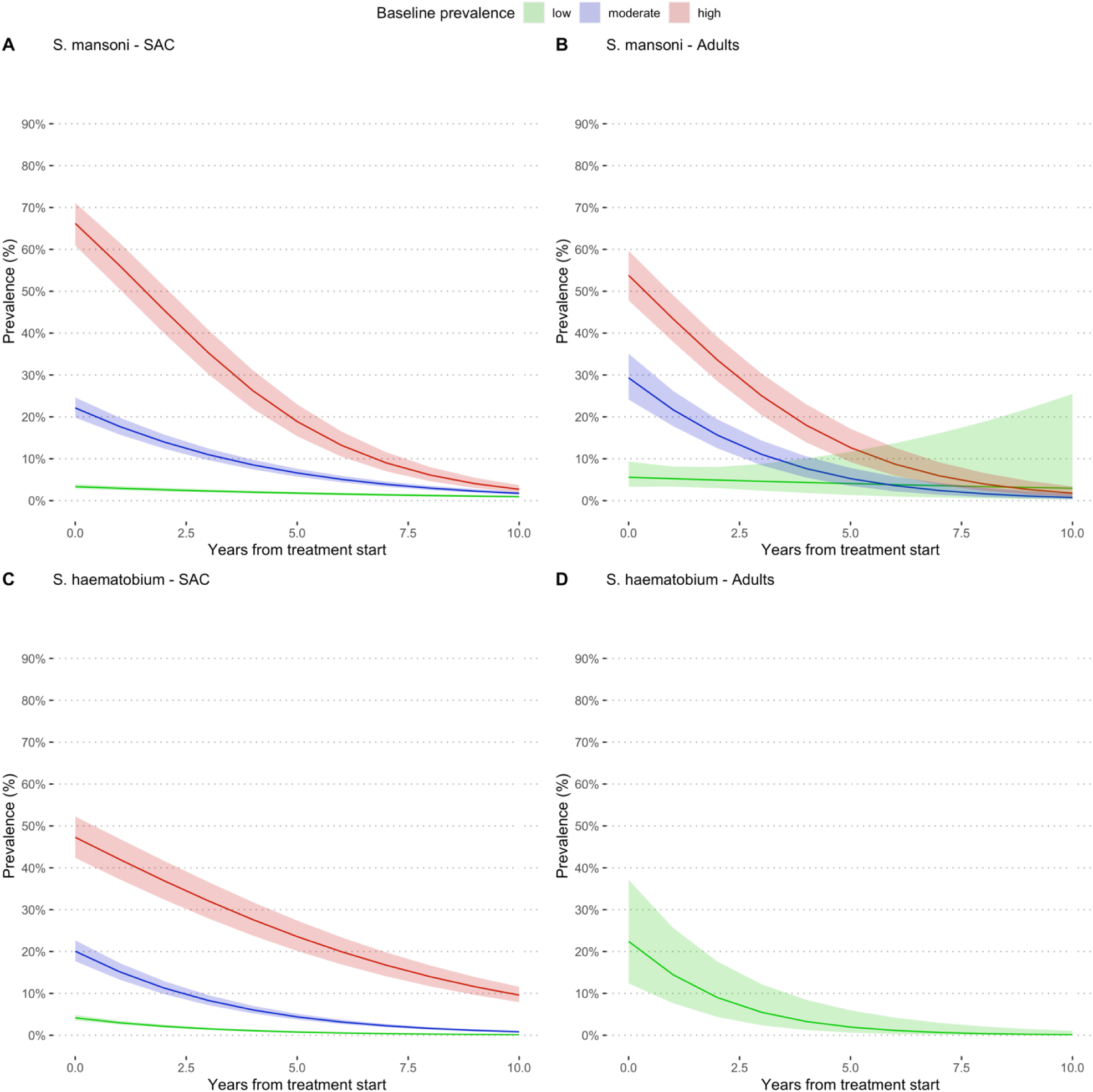
Estimated prevalence at baseline and at different follow-up times during MDA for *S. mansoni* (A-B) and *S. haematobium* (C-D). High prevalence (pink colour) is defined as prevalence ≥50% by microscopy and ≥60% by CCA, moderate (blue colour) is prevalence =10% but <50% by microscopy and ≤ 15% but <60% by CCA and low prevalence (green colour) is prevalence <10% by and <15% by CCA. All the studies used PZQ 40 mg/kg administered as a single oral dose alone or in combination with Albendazole (400 mg) in MDA campaigns in endemic countries in Sub-Saharan Africa.

**Fig. 9.**
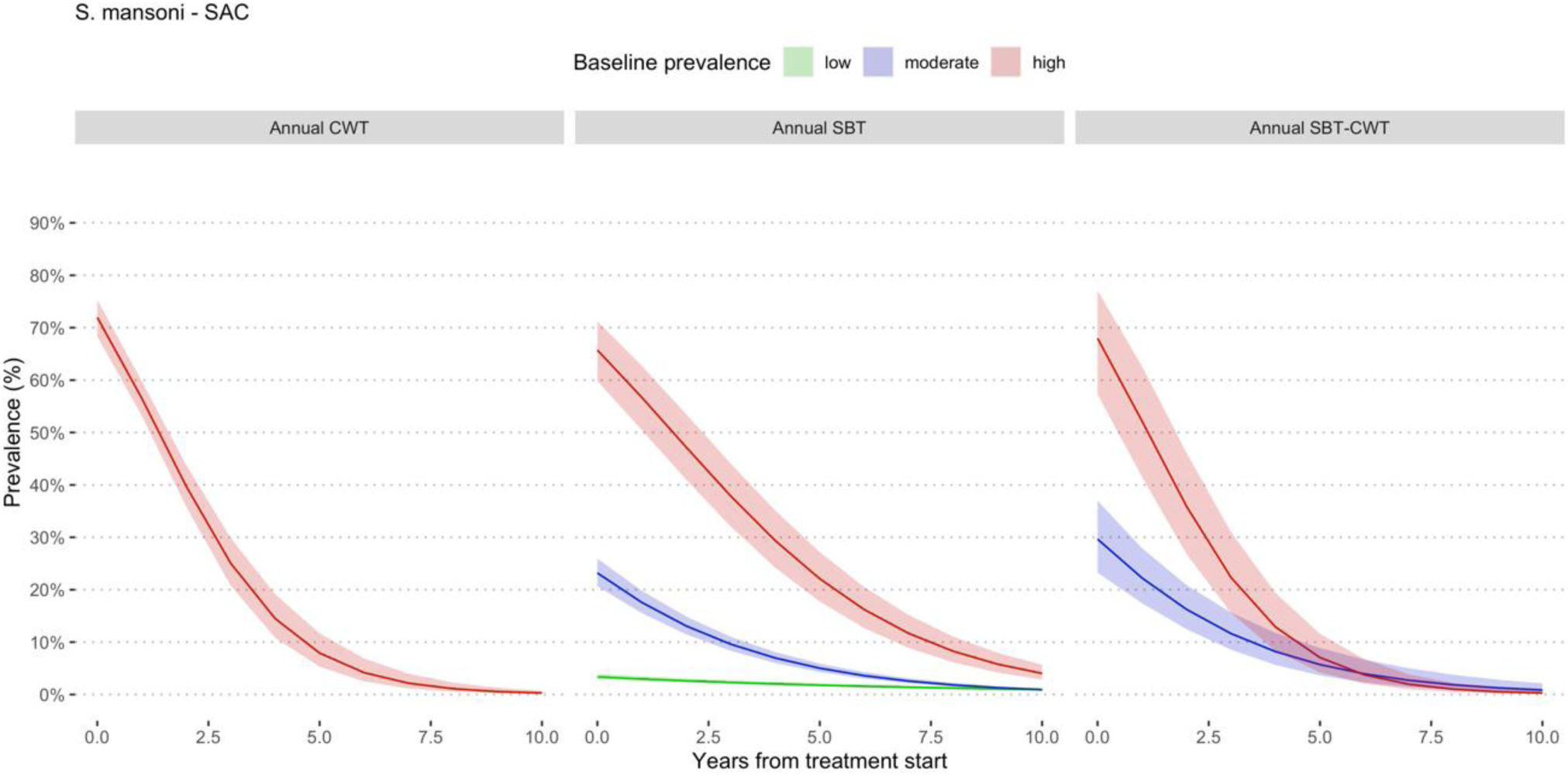
Estimated prevalence at baseline and at different follow-up times during MDA involving school-age children for *S. mansoni*. Prevalence categories have been defined using WHO classification.^6^ High prevalence is defined as prevalence ≥50% by microscopy and ≥60% by CCA, moderate is prevalence =10% but <50% by microscopy and ≤ 15% but <60% by CCA and low prevalence is prevalence <10% by and <15% by CCA. All the studies used PZQ 40 mg/kg administered as a single oral dose alone or in combination with Albendazole (400 mg) in MDA campaigns in endemic countries in Sub-Saharan Africa. SBT means school-based treatment and CWT community-wide treatment.

Further theory-grounded analyses, supported by the biology and epidemiology of schistosomiasis, created prevalence thresholds at 5%, 10%, 15%, 20%, 30% and ≥40% to determine the most operationally feasible and cost-effective threshold to guide the implementation of MDA programmes (Fig. 10 and 11). The different thresholds showed differences in effectiveness when used to guide the implementation of MDA programmes, with 10% threshold demonstrating best outcome (Fig. 10). It took around 10 years to reduce prevalence to 1% with repeated annual MDA, whereas for higher thresholds (15%, 20%, 30% and ≥40%), it will take up to 15 years for prevalence to reduce to 1% from extrapolated data. The graphs show that after decreasing to 1%, repeated MDA will have minimal effect in bringing prevalence down further to 0%. For MDA delivered using CWT strategy for SAC in *S. mansoni* endemic areas, the data showed that at 10% threshold, repeated annual CWT was unable to reduce prevalence to 1% after 10 years of repeated MDA. The extrapolated model suggests it will take up to 15 years to achieve this; higher thresholds though will take longer time than 15 years to reduce prevalence to 1%. Again, it appears it will take a considerable effort to push prevalence down beyond 1% to 0% with repeated annual MDA. The results were similar for *S. haematobium* with immediate impact of MDA, demonstrated by a sharp drop of prevalence, particularly with 30% and ≥40% thresholds (Fig. 11). The drop is steeper for SBT than CWT suggesting, perhaps, better immediate impact of SBT than CWT. For all thresholds, SBT MDA for SAC was able to reduce prevalence to 5% around 5 years and 1% up to 10 years. In fact, 5% threshold was able to reduce prevalence of *S. haematobium* to 0% around 10 years, but it will take longer time to achieve this with 10% or 15% threshold. For annual CWT, the curves show that 20%, 15% and 5% are doing better but not 10% threshold, although none achieved a decrease to 1% by 10 years. The shapes of the curves suggest that it will take considerable effort and time to reduce prevalence further beyond 1% with annual MDA; it may take additional 5 to 10 years to bring prevalence to 0%. From the pooled overall data in this meta-analysis, 10% threshold (for *S. mansoni*) and 5% threshold (for *S. haematobium*) implemented through SBT of SAC and adults appear to be the most effective strategy for controlling schistosomiasis within the PC concept.

**Fig 10.**
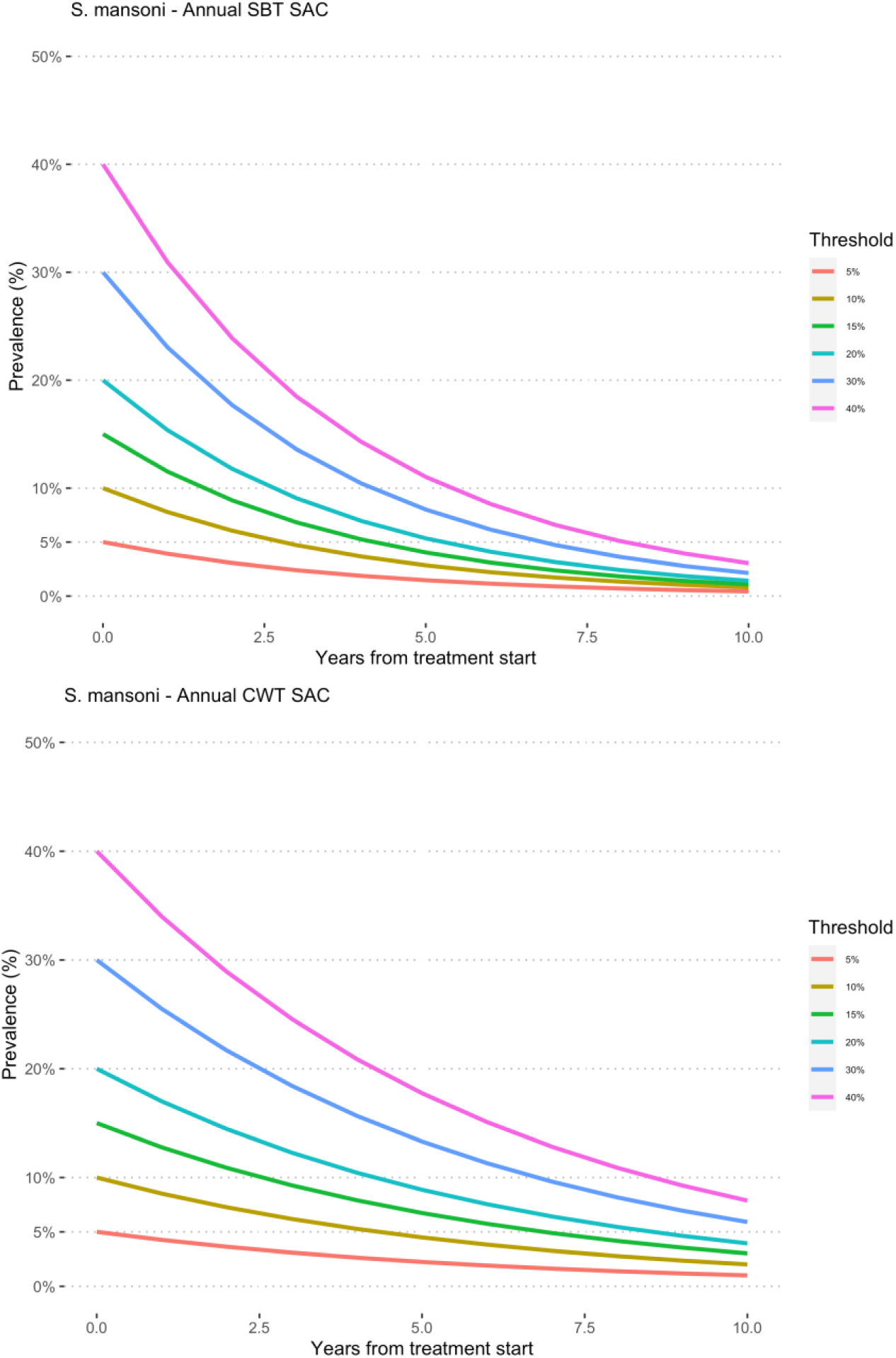
Prevalence rate reduction in SAC MDAs for *S. mansoni* infection according to longitudinal analyses using different thresholds created from the data generated from this systematic review, with 5% representing the lowest prevalence threshold, followed by 10%, 15%, 20%, 30% and 40% thresholds. For a particular threshold, the corresponding reduction in prevalence over time after SBT (School-Based Treatment) and CWT (community-wide treatment) MDA are represented by the different contours.

**Fig. 11.**
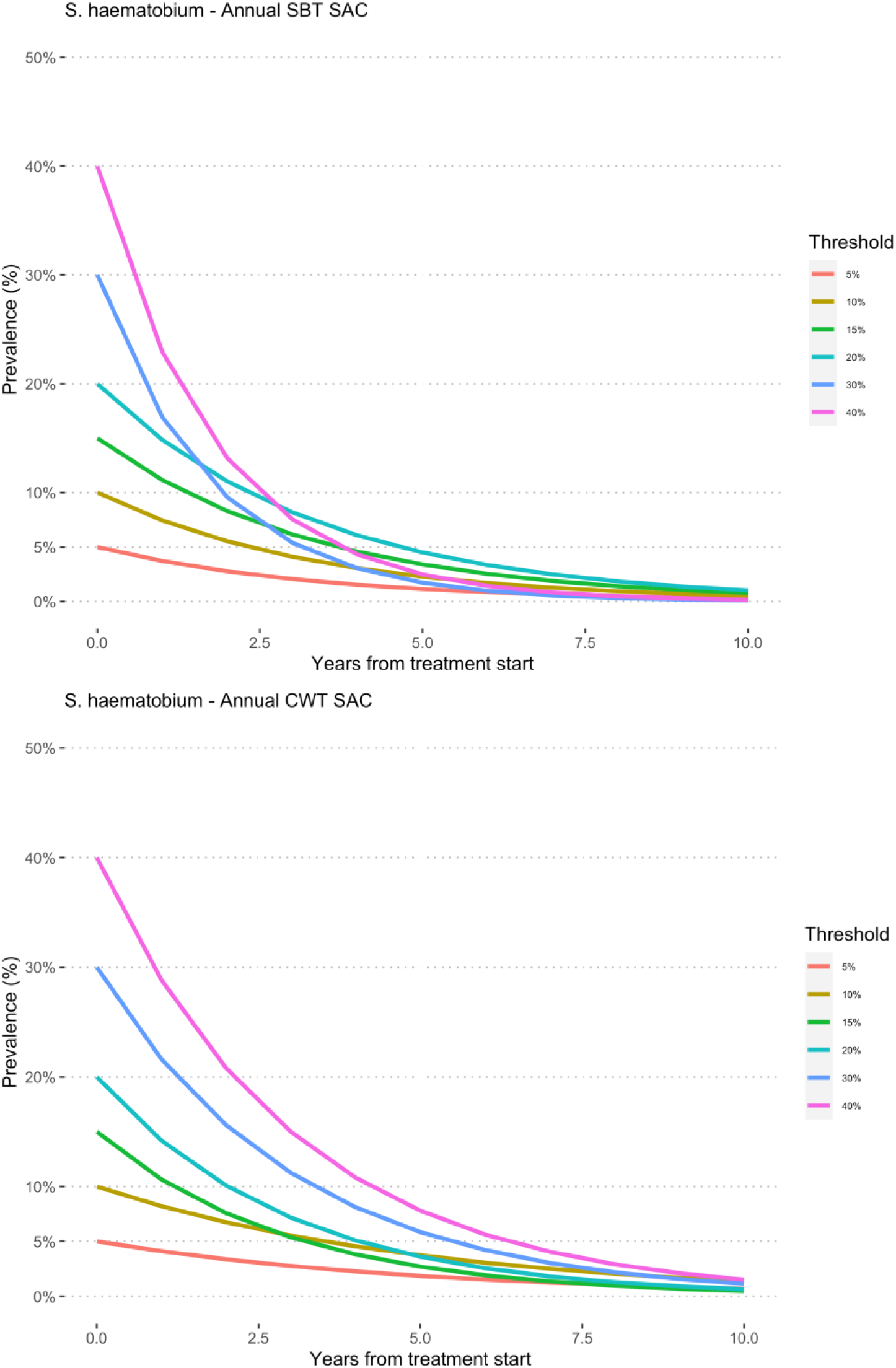
Prevalence rate reduction in SAC MDAs for *S. haematobium* infection according to longitudinal analyses using different thresholds from data generated by this systematic review, with 5% representing the lowest prevalence threshold, followed by 10%, 15%, 20%, 30% and 40% thresholds. For a particular threshold, the corresponding reduction in prevalence over time after SBT and CWT are represented by the different contours.

### Risk of Bias and level evidence

The risk of bias in the included studies is represented graphically (Fig. S4). The overall risk of bias in the 34 studies included the meta-analysis, which the majority were observational studies, was moderate. All the studies were considered as low Risk of Bias selection of the reported results, deviations from intended interventions, classification of interventions, selection of participants into the study and confounding. For measurement of outcomes, all the studies were considered as moderate risk of bias whereas only three studies had information on missing data and were scored as moderate, the rest (31 studies) did not report on missing data and were rated as NI (no information). The risk of bias scores for the individual studies have been reported in Table S4. The overall quality of evidence as assessed using GRADE was moderate (Table 4).

**Table 4.** Level of evidence using GRADE (Grading of Recommendations, Assessment, Development and Evaluations)

| Anticipated absolute effects*<br>(95% CI) |  |  |  |  |  |
| --- | --- | --- | --- | --- | --- |
| Outcomes | Without preventive chemotherapy | With preventive chemotherapy | Relative effect (95% CI) | Number of participants (studies) | Quality of the evidence (GRADE) |
| One-year prevalence reduction of *S. mansoni* in SAC (Annual SBT; 0 to 1 year) | 8,155/45,510 (17.9%) | 5,047/40,563 (12.4%) | PR = 0.56 (0.46 to 0.69) | 45,510 (14) | ⊕⊕⊕⊙ |
| 4 years prevalence reduction of *S. mansoni* in SAC (Annual SBT; 0 to 4 year) | 3,936/17,745 (22.2%) | 1,196 /9,738 (12.3%) | PR = 0.25 (0.11 to 0.59) | 17,745 (5) | ⊕⊕⊕⊙ |
| Annual prevalence reduction of *S. mansoni* in SAC (Annual SBT; Threshold >5%; follow-up: range 1-7 years) | - | - | PR = 0.78 (0.77 to 0.79) | 123,045 (20) | ⊕⊕⊕⊙ |
| One-year prevalence reduction of *S. haematobium* in SAC (Annual SBT; 0 to 1 year) | 6,582/20,040 (32.8%) | 3,027/17,828 (17.0%) | PR = 0.38 (0.28 to 0.52) | 20,040 (8) | ⊕⊕⊕⊙ |
| 4 years prevalence reduction of *S. haematobium* in SAC (Annual SBT; 0 to 4 years) | 1,387/6,274 (22.1%) | 949/4,680 (20.3%) | PR = 0.38 (0.28 to 0.52) | 6,724 (2) | ⊕⊕⊙⊙ |
| Annual prevalence reduction of *S. haematobium* in SAC (Annual SBT; Threshold >5%; follow-up: range 1-7 years) | - | - | PR = 0.74 (0.73 to 0.75) | 106,912 (13) | ⊕⊕⊕⊙ |

GRADE Working Group grades of evidence (Guyatt et al., 2013)
⊕⊕⊕⊕ High quality: We are very confident that the true effect lies close to that of the estimate of effect.
⊕⊕⊕⊙ Moderate quality: We are moderately confident in the effect estimate: The true effect is likely to be close to the estimate of the effect, but there is a possibility that it is substantially different.
⊕⊕⊙⊙ Low quality: Our confidence in the effect estimate is limited: The true effect may be substantially different from the estimate of the effect.
⊕⊙⊙⊙ Very low quality: We have very little confidence in the effect estimate: The true effect is likely to be substantially different from the estimate of effect.

## Discussion

### Summary of the main results

The results of the systematic review and meta-analysis showed there is no direct correlation between prevalence and intensity of schistosome infections. Praziquantel reduced prevalence of *S. mansoni* infection and sustained the reduction in school age children following mass drug administration for up to 48 months where this was slightly shorter for *S. haematobium* (up to 36 months). The optimal prevalence threshold that will lead to the most cost-effective preventive chemotherapy programmes was estimated to be 10% for both *S. mansoni* and *S. haematobium*. Repeated annual MDA delivered as school-based treatment showed to be the most effective strategy of delivering preventive chemotherapy. Using regression models, it was estimated that when using the optimal 10% prevalence threshold, it will take over 10 years to reduce prevalence of *S. haematobium* to 1% and up to 15 years for *S. mansoni* infection. The overall pooled data suggests that at the 10% threshold, school-based treatment of SAC plus adults will be the most effective mass drug administration strategy for controlling schistosomiasis withing the PC concept.

There was a sharp drop in prevalence rate after MDA but the decrease became less prominent with the curves appearing to be leveling off over time, particularly for *S. mansoni*. This may have implications for control as it suggests that prevalence can drop after MDA but at some point, it becomes difficult to reduce prevalence further by repeated MDA.

Given the evidence presented that showed 10% prevalence threshold will achieve elimination with community wide MDA at around 10 years, coupled with an average baseline prevalence of most endemic settings where MDA is required is around 17% (assuming a margin of error of 7%; almost all settings will be covered) and the fact that higher threshold will take up to 15 years to achieve elimination, 10% prevalence threshold is recommended as universal threshold to implement MDA in endemic settings.

Heterogeneity between the studies pooled in this meta-analysis was high (*I^2^ >* 95% in all the analysis) but given the huge number of studies and individuals treated, between studies heterogeneity is not expected to change the direction of treatment effect or the overall conclusions from this review. The high levels of heterogeneity could be explained by the following: 1) there are ‘real’ differences between MDA programmes so the *I^2^* values are a true reflection, 2) MDA programmes differed in strategy (some treated only SAC, SAC and occupationally at risk adults (SAC and adults) or community wide treatment, 3) other control measures took place during the period of the MDA in some cases, 4) villages differed in baseline intensity of infection and thereby force of transmission, 5) variation in sensitivities in diagnostic criteria existed between MDA programmes, and 6) differences existed in follow-up times of MDAs. Data on some of these factors were not documented or presented in formats that could be used in this systematic review and meta-analysis and, therefore, some of these variations could not be quantitatively addressed in the meta-analysis or in sub-group analyses. Integration of non-pharmacological interventions such access to water, improved sanitation, hygiene education (WASH) and snail control will complement MDA to achieve elimination of schistosomiasis.

### Agreements and disagreements with other studies or reviews

From the meta-regression model, prevalence and intensity of infection showed nonlinear, but quadratic polynomial relationship. This has implications for control using prevalence threshold derived from egg count or CCA as high prevalence may not necessarily translate into high intensity of infection and vice versa. Although some studies have assumed a linear relationship between prevalence and intensity if infection, others have suggested a nonlinear relationship. Some authors have even gone further to suggest that intensity is more stable than prevalence and can best predict the success of control over a long period of time. Given that the assumed linear relationship held by many appears not to be accurate, at least from evidence generated in this systematic review and meta-analysis, caution is needed when interpretating and justifying the use of prevalence threshold to guide the implementation of an MDA. There is a need for more work to determine an optimal threshold based on intensity of infection so both prevalence and intensity thresholds can be compared to ascertain which one is more stable and responsive MDA across varying settings and contexts, and also robust to predict long term progress and success of MDA programmes.

### Overall completeness and applicability of Evidence

GRADE (Grading of Recommendations, Assessment, Development and Evaluations) is a transparent framework for developing and presenting summaries of evidence and provides a systematic approach for making clinical practice recommendations [1-3] It is the most widely adopted tool for grading the quality of evidence and for making recommendations with over 100 organisations worldwide officially endorsing GRADE.

The results from this study showed that annual MDA leads to prevalence reduction of up to 48 months in school-age children with schistosomiasis, but the effect levels off at 36months for *S. haematobium* and 48 months for *S. mansoni* although the evidence is inconclusive as only two studies were involved at 48 months. Strikingly, there were no apparent further reductions in prevalence with repeat annual MDA over that attained at 12 months for both *S. haematobium* and *S. mansoni*. The community-wide treatment strategy showed similar results. This finding is supported by a series of studies conducted in endemic settings (Ouattara et al. 2021). The implications for this are serious, as it suggests that although there are benefits, MDA alone is not likely to achieve elimination in endemic countries.

For example, annual mass drug administration for up to 9 years did not achieve prevalence reduction at the elimination target for *S. mansoni* in settings with baseline prevalence ≥10%. For *S. haematobium*, although annual mass drug administration up to 5 years reduced prevalence to lower levels than *S. mansoni,* elimination target was not reached in setting with baseline prevalence ≥10% by 10 years of MDA. From extrapolated analysis up to 15 years of repeated annual whole community MDA will be required to achieve schistosomiasis elimination when baseline prevalence is ≥10%. Further analyses using a series of thresholds created from the data, no significant differences in the times to elimination were observed between 5% and 10%, 15%, 20%, 30% and 40%higher than 10% threshold took longer to reach elimination.

Heterogeneity between the studies pooled in this meta-analysis was high, reaching *I^2^* >97%. However, but most of the studies had similar characteristics in terms of population, intervention and outcomes, and measured the same outcomes. The major difference was the degrees of baseline endemicity which varied across studies, ranging from …to…. . Another major issue relates to the strategies of MDA employed where some MDA programmes treated only SAC (sbt), SAC and occupationally at-risk adults (SAC and adults) or community-wide treatment (cwt). In some settings, other control measures took place during the same period of the MDA, villages differed in baseline intensity of infection and thereby force of transmission, variation in sensitivities in diagnostic criteria existed between MDA programmes as well as differences in follow-up times after MDA. Mode of delivery of MDA differed between studies: majority of studies delivered annual MDA for at least two years, some studies delivered MDA twice a year (bi-annual) and for some studies, MDA was delivered every two years (biennial). Some studies delivered annual MDA for two years and assessed effect of the treatment after 5 years so that three years elapsed after the last MDA before the outcome of treatment was assessed and some studies alternated MDA with drug holidays. As data on some of these factors were not documented or in formats that could be used, these variations could not be quantitatively addressed in the meta-analysis or sub-group analysis. This has exposed the lack of standardization across MDA programmes and the implications for schistosomiasis control. We suggest future research should use standardised tools, methods and approach in order to reduce variation across control programmes. However, the fact that the number of individuals in the included studies were very high, it is not likely that the study conclusions could have been influenced by heterogeneity alone.

The high levels of heterogeneity could be explained by the following: 1) there are ‘real’ differences between MDA programmes, so the *I^2^* values are a true reflection, 2) MDA programmes differed in strategy (some treated only SAC, SAC and occupationally at-risk adults (SAC and adults) or community-wide treatment, 3) other control measures took place during the period of the MDA in some cases but not accounted for in reporting the results of MDAs, 4) villages differed in baseline intensity of infection and thereby force of transmission, 5) variation in sensitivities in diagnostic criteria existed between MDA programmes, and 6) differences existed in follow-up times of MDAs.

Data on some of these factors were not documented or presented in formats that could be used in this systematic review and meta-analysis, and therefore, some of these variations could not be quantitatively addressed in the meta-analysis or in sub-group analyses. Most of the data are from SAC population; no data on pre-school children and few on adults make it difficult to make a concrete statement about the generalizability of the review findings beyond SAC. From this review, PZQ appears to be effective in reducing the prevalence of schistosomiasis at 12 months, but the incremental benefit of repeated annual treatment appears to be minimal in further reducing prevalence over time. In both light prevalence and intensity, infection rates do not decrease but rather tended to increase with biennial MDA. Effectiveness depends on several factors, which are difficult to disentangle. However, the rate of prevalence decrease seems not influenced by the baseline intensity of infection and type of treatment. Further analyses were conducted from a series of created prevalence thresholds of 5%, 10%, 15%, 20%, 30% and ≥40% and although showed differences in the effect of MDA, the differences in effect based on threshold regressed over time (after about 8 years). For the annual MDA of SAC, SBT appears to perform better than CWT in terms of prevalence reduction. For schistosome species, the model suggests, using the same prevalence threshold, it will take shorter time to reach elimination for *S. haematobium* than *S. mansoni*; annual MDA for *S. haematobium* (SBT) will require about 10 years to achieve elimination whereas it will take longer for *S. mansoni* (over 10 years to around 15 years to achieve elimination). The question that remains is what happens to endemic communities/settings with less than 10% prevalence? Integration of non-pharmacological interventions such as access to water, improved sanitation, hygiene education (WASH) and snail control will complement MDA to achieve the elimination of schistosomiasis.

### Conclusions and recommendations

The evidence presented in this review demonstrates that 10% is the optimum prevalence that should be used as the global prevalence threshold for deciding PC campaigns in endemic countries.

### Implications for practice Implications for policy Implications for research

We suggest a head-to-head comparison of prevalence and intensity thresholds to ascertain which of the two is more stable and responsive MDA across settings and contexts.

## Data Availability

This review was commissioned by the World Health Organization (WHO) and data will be made available to the public.

## Additional references

Mamadou Ouattara, Nana R Diakité, Patrick K Yao, Jasmina Saric, Jean T Coulibaly, Rufin K Assaré, Fidèle K Bassa, Naférima Koné, Négnorogo Guindo-Coulibaly, Jan Hattendorf, Jürg Utzinger, Eliézer K N’Goran. Effectiveness of school-based preventive chemotherapy strategies for sustaining the control of schistosomiasis in Côte d’Ivoire: Results of a 5-year cluster randomized trial. PLoS Negl Trop Dis. 2021 Jan 15;15(1):e0008845. doi: 10.1371/journal.pntd.0008845. eCollection 2021 Jan.

## APPENDIX 1

**Table S1.**
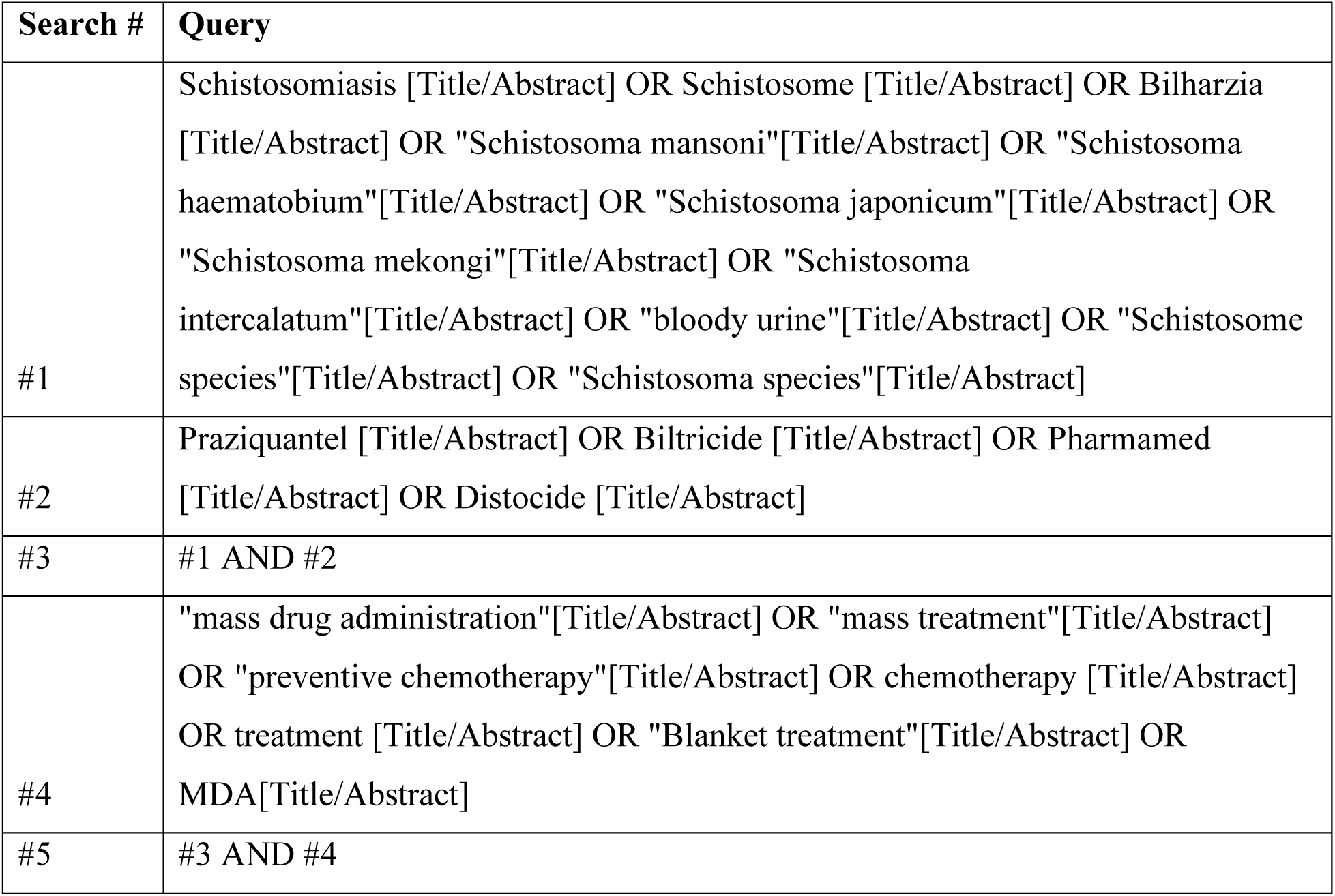
Search strategy and search terms

**Table S2.**
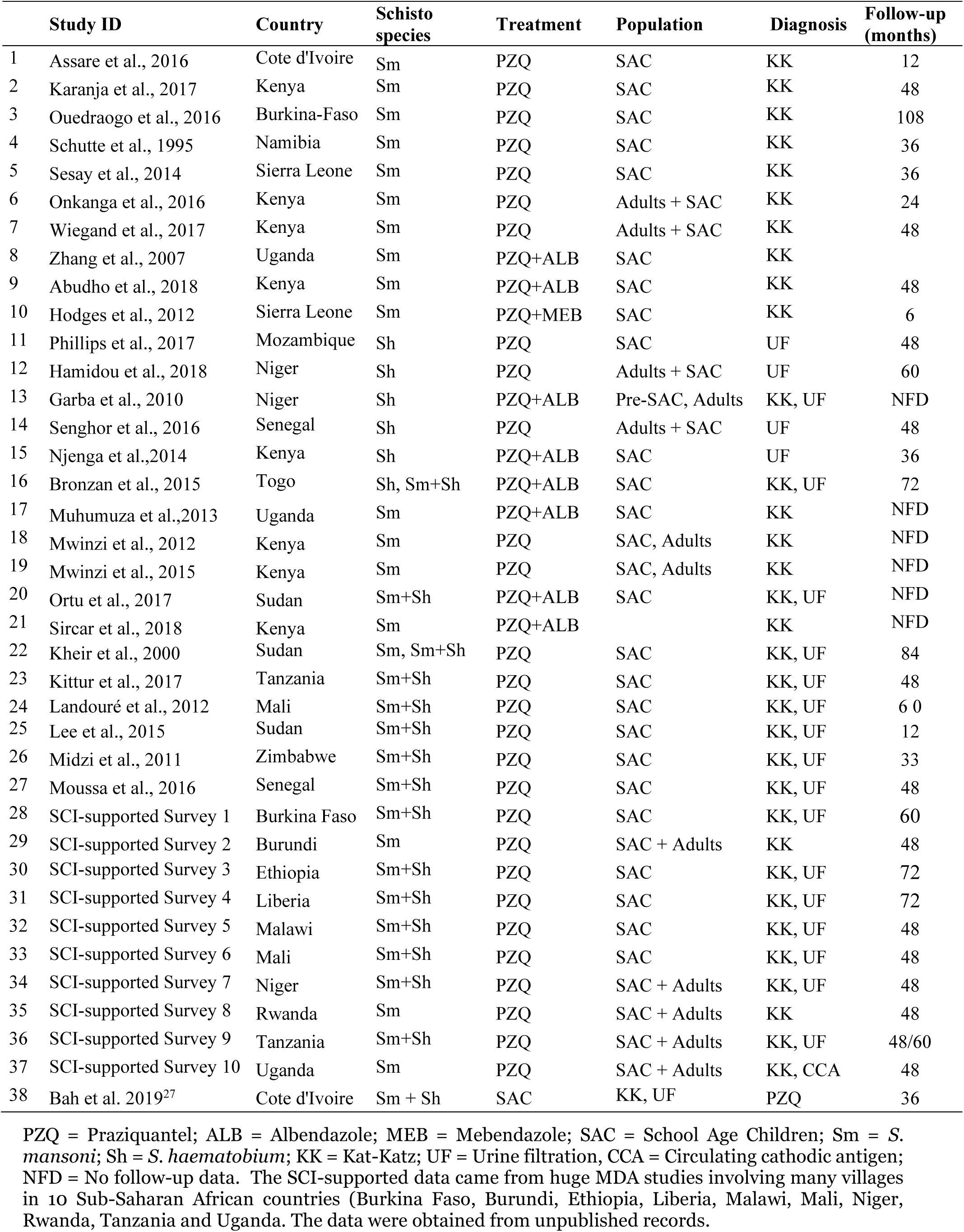
Characteristics of the studies meeting the inclusion criteria

**Table S3.** Meta-regression results in terms of Prevalence Ratios (95% CI) per year of treatment*

|  | Studies | PR (95% CI) | p-value |
| --- | --- | --- | --- |
| *S. mansoni* | 28 | 0.827 (0.823 to 0.832) | <0.001 |
| *S. haematobium* | 19 | 0.756 (0.751 to 0.761) | <0.001 |

\*Meta-analyses were performed separately for each species. Estimates are derived from GLLM for longitudinal data.

**Fig. S2b.**
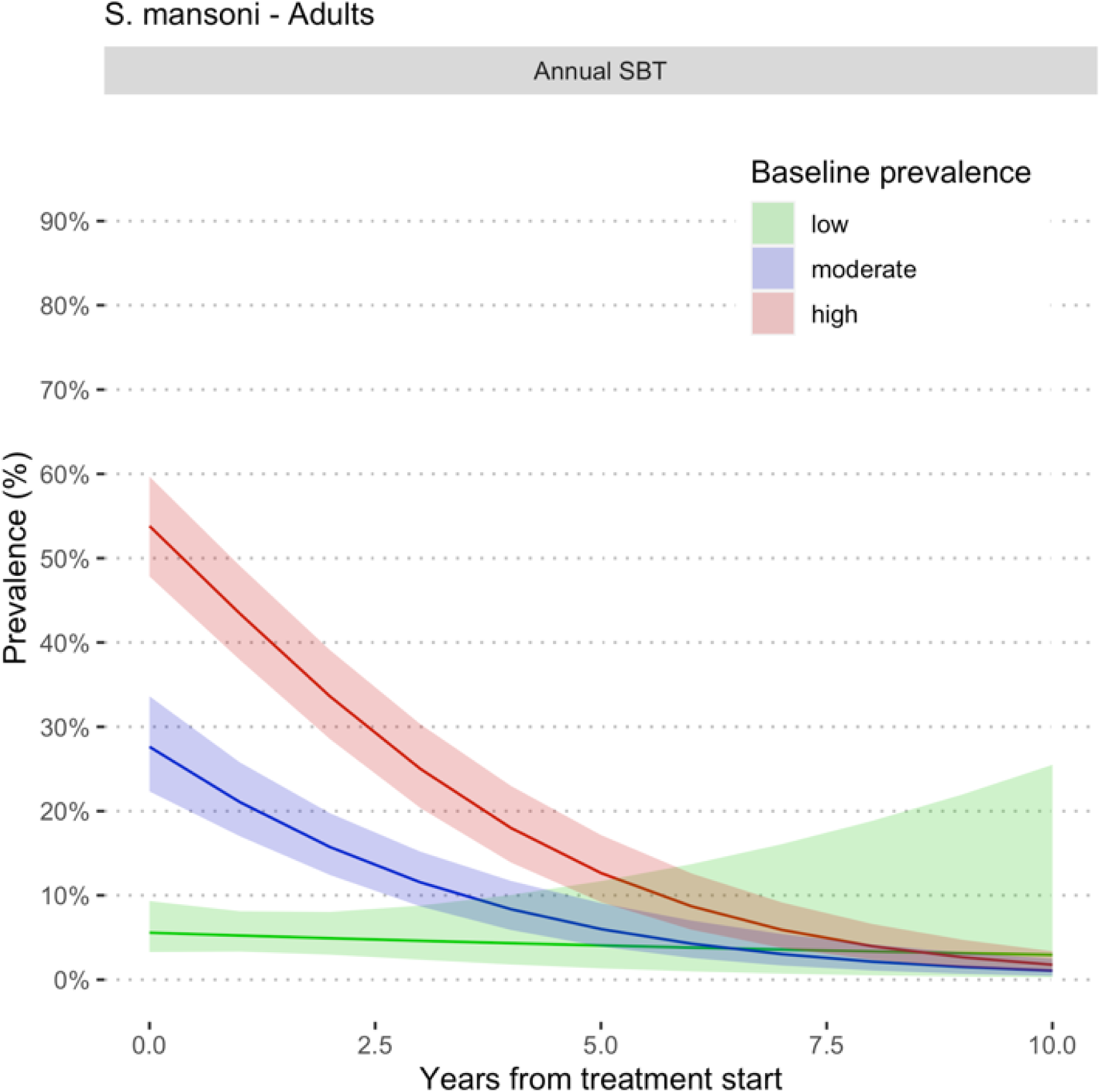
Estimated prevalence at baseline and at different follow-up times during MDA involving Adults for *S. mansoni*. Prevalence categories have been defined using WHO classification.^6^ Low prevalence (green colour) is prevalence <10% when using microscopy for infection detection and <15% by CCA, moderate (blue colour) is prevalence =10% but <50% by microscopy and ≤ 15% but <60% by CCA and high prevalence (pink colour) is prevalence ≥50% by microscopy and ≥60% by CCA. All the studies used PZQ 40 mg/kg administered as a single oral dose alone or in combination with Albendazole (400 mg) in MDA campaigns in endemic countries in Sub-Saharan Africa. SBT means school-based treatment.

**Fig. S2c.**
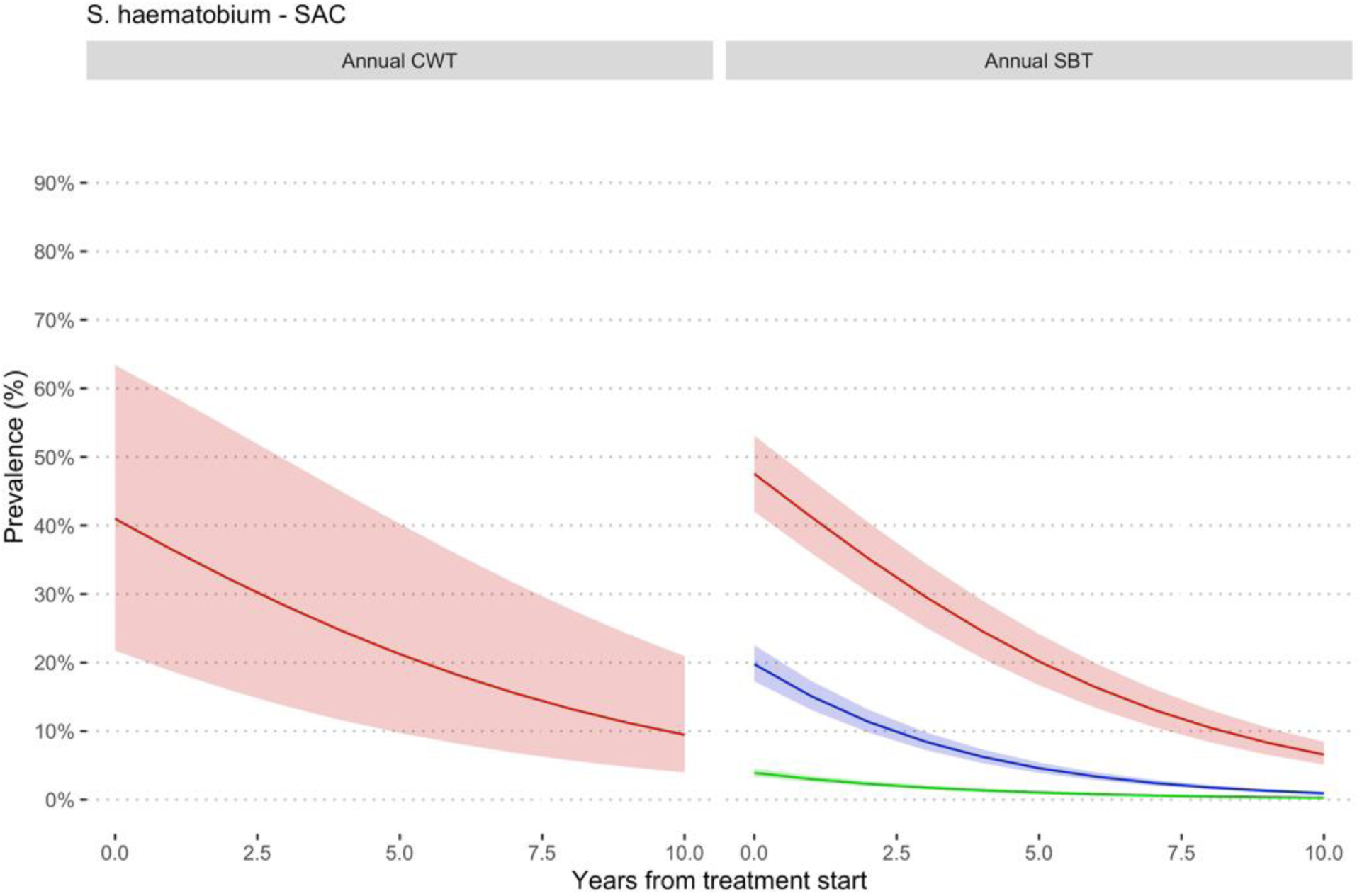
Estimated prevalence at baseline and at different follow-up times during MDA involving school-age children for *S. haematobium*. Prevalence categories have been defined using WHO classification.^6^ Low prevalence (green colour) is prevalence <10% when using microscopy for infection detection and <15% by CCA, moderate (blue colour) is prevalence =10% but <50% by microscopy and ≤ 15% but <60% by CCA and high prevalence (pink colour) is prevalence ≥50% by microscopy and ≥60% by CCA.All the studies used PZQ 40 mg/kg administered as a single oral dose alone or in combination with Albendazole (400 mg) in MDA campaigns in endemic countries in Sub-Saharan Africa. SBT means school-based treatment and CWT community-wide treatment.

**Fig. S2d.**
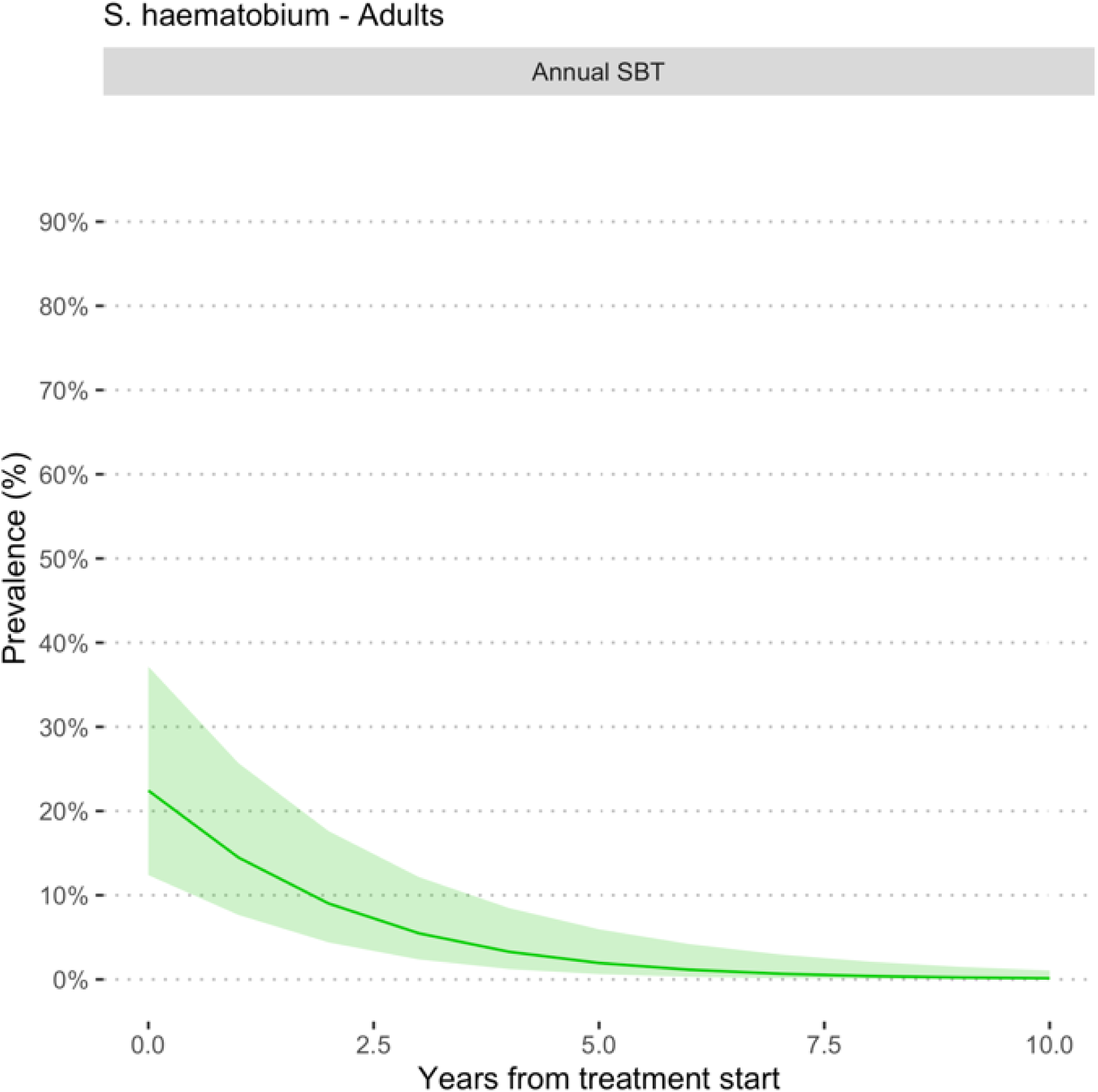
Estimated prevalence at baseline and at different follow-up times during MDA involving Adults for *S. haematobium*. Prevalence categories have been defined using WHO classification.^6^ High prevalence is defined as prevalence ≥50% by microscopy and ≥60% by CCA, moderate is prevalence =10% but <50% by microscopy and ≤ 15% but <60% by CCA and low prevalence is prevalence <10% by and <15% by CCA. All the studies used PZQ 40 mg/kg administered as a single oral dose alone or in combination with Albendazole (400 mg) in MDA campaigns in endemic countries in Sub-Saharan Africa. SBT means school-based treatment.

**Fig. S3a.**
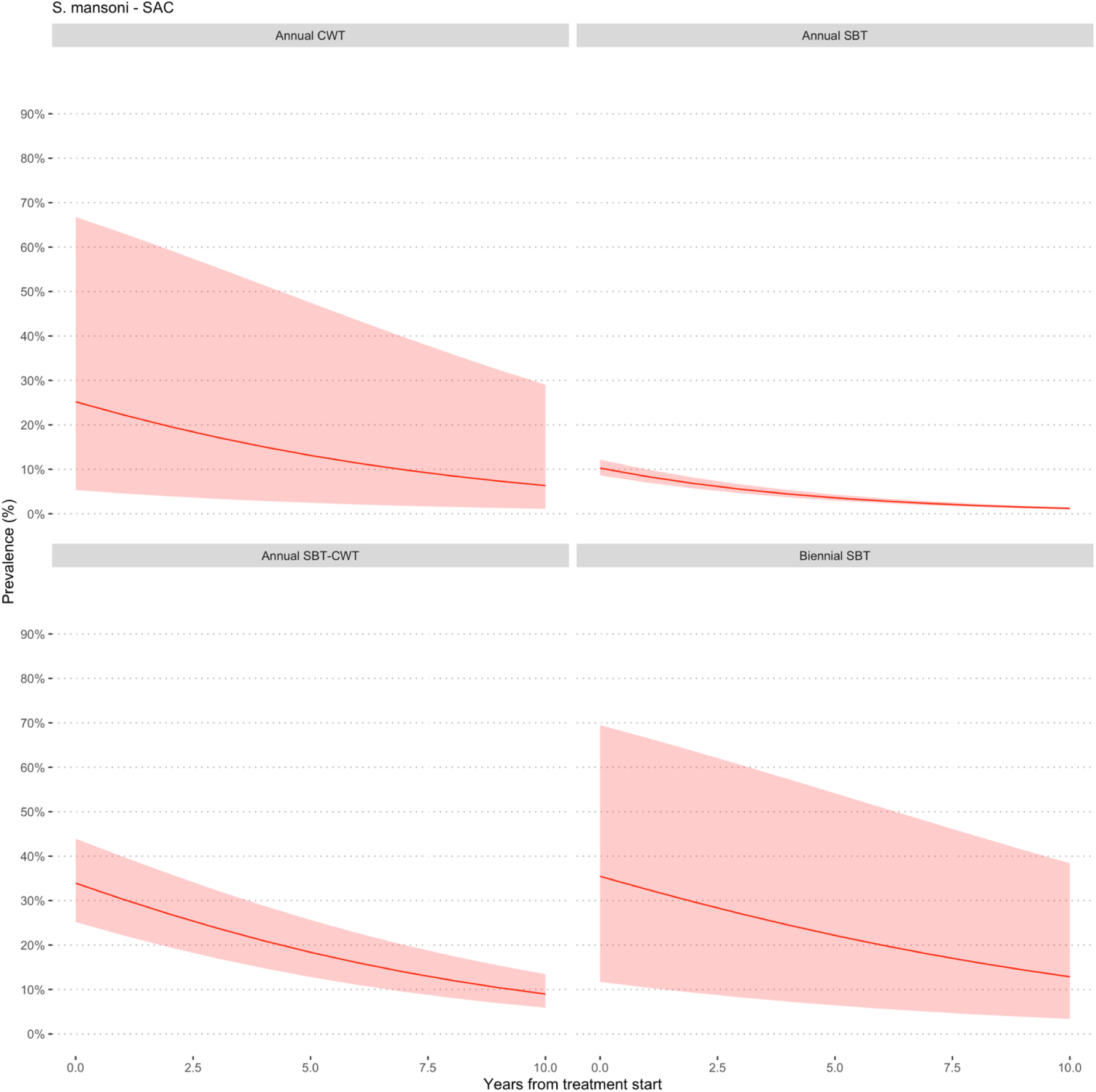
Estimated prevalence at baseline and at different follow-up times during MDA involving school-age children for *S. mansoni*. All the studies used PZQ 40 mg/kg administered as a single oral dose alone or in combination with Albendazole (400 mg) in MDA campaigns in endemic countries in Sub-Saharan Africa.

**Fig. S3b.**
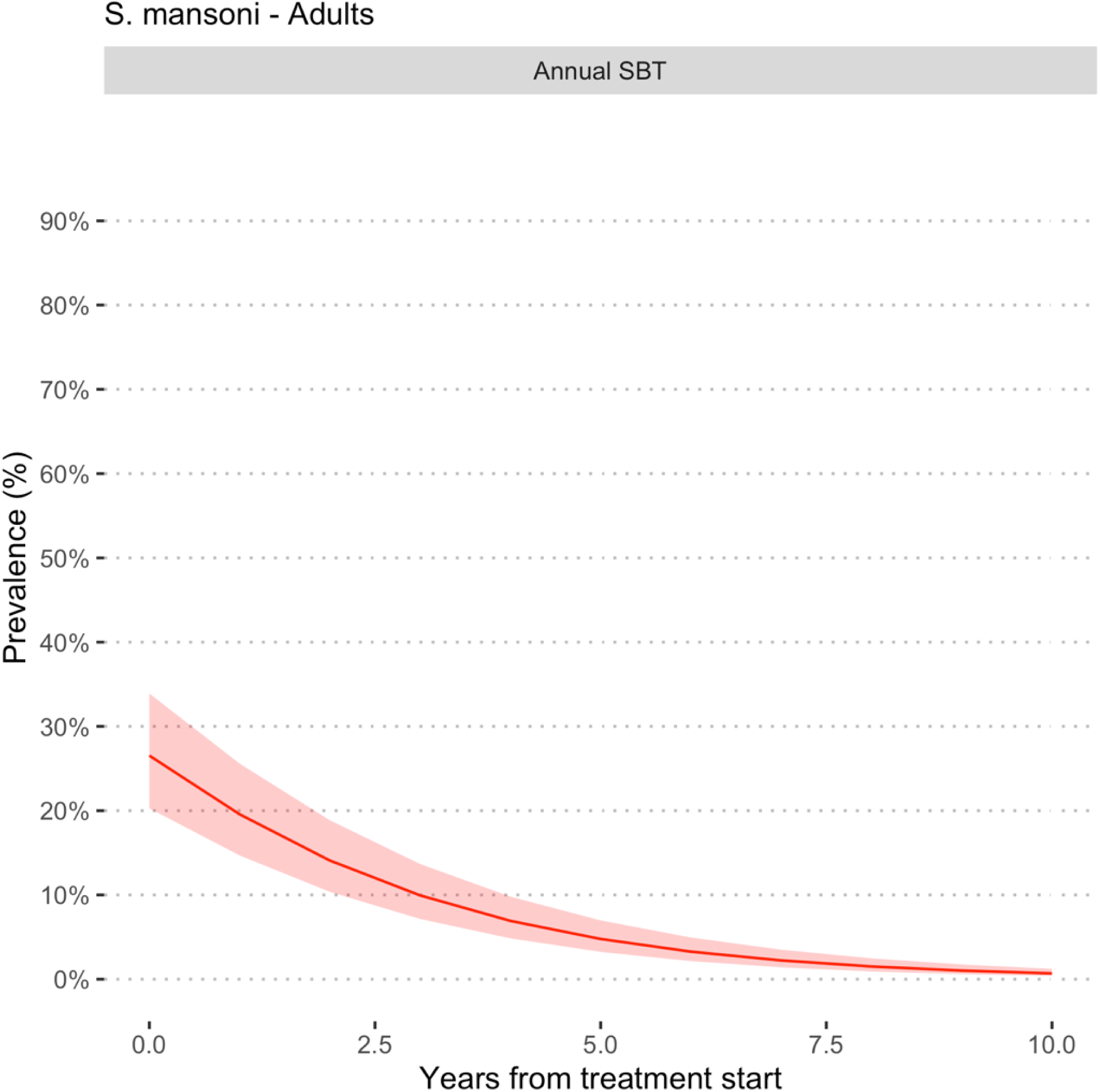
Estimated prevalence at baseline and at different follow-up times during MDA involving adults for *S. mansoni*. All the studies used PZQ 40 mg/kg administered as a single oral dose alone or in combination with Albendazole (400 mg) in MDA campaigns in endemic countries in Sub-Saharan Africa.

**Fig. S3c.**
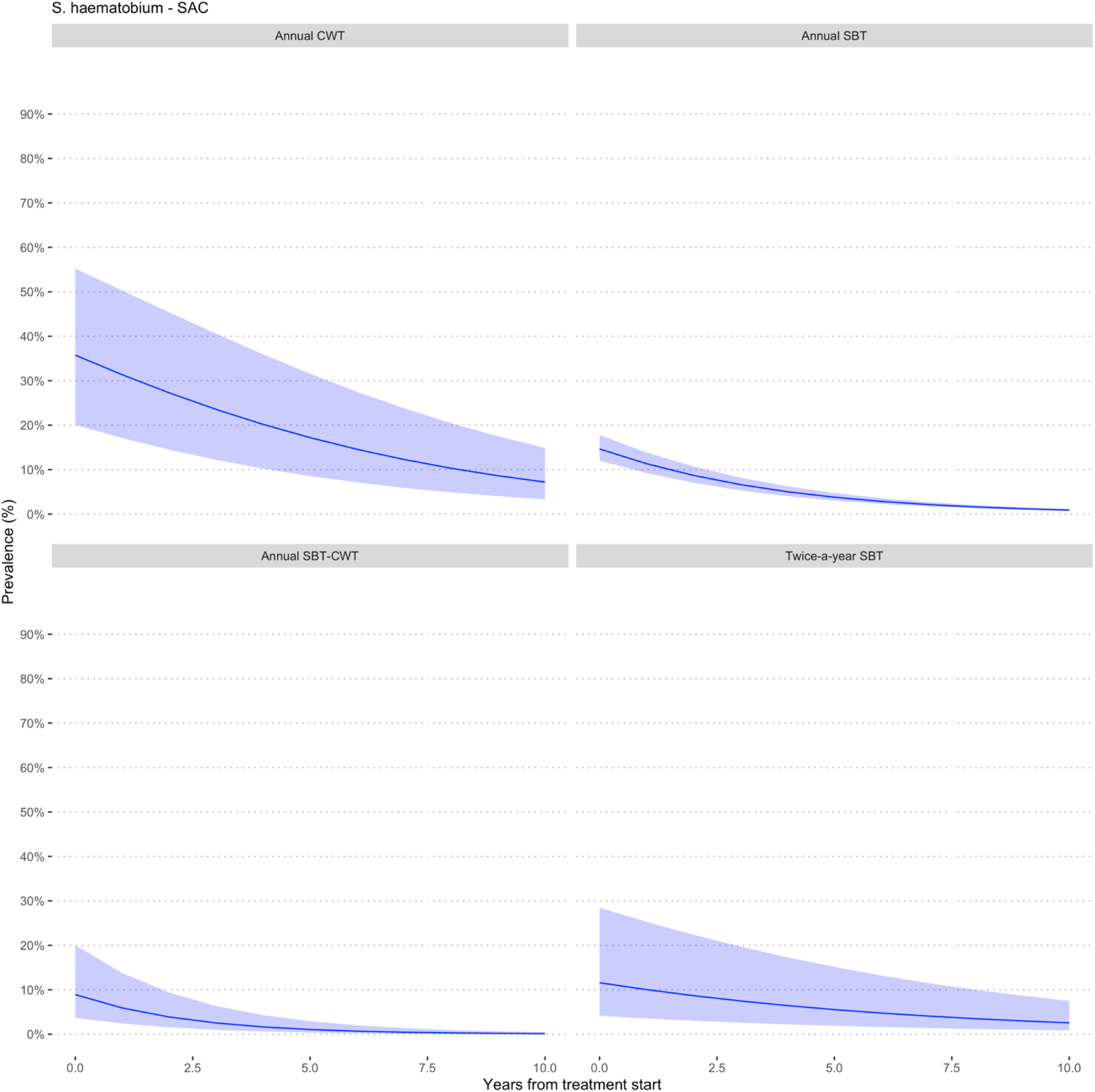
Estimated prevalence at baseline and at different follow-up times during MDA involving school-age children for *S. haematobium*. All the studies used PZQ 40 mg/kg administered as a single oral dose alone or in combination with Albendazole (400 mg) in MDA campaigns in endemic countries in Sub-Saharan Africa.

**Fig. S3d.**
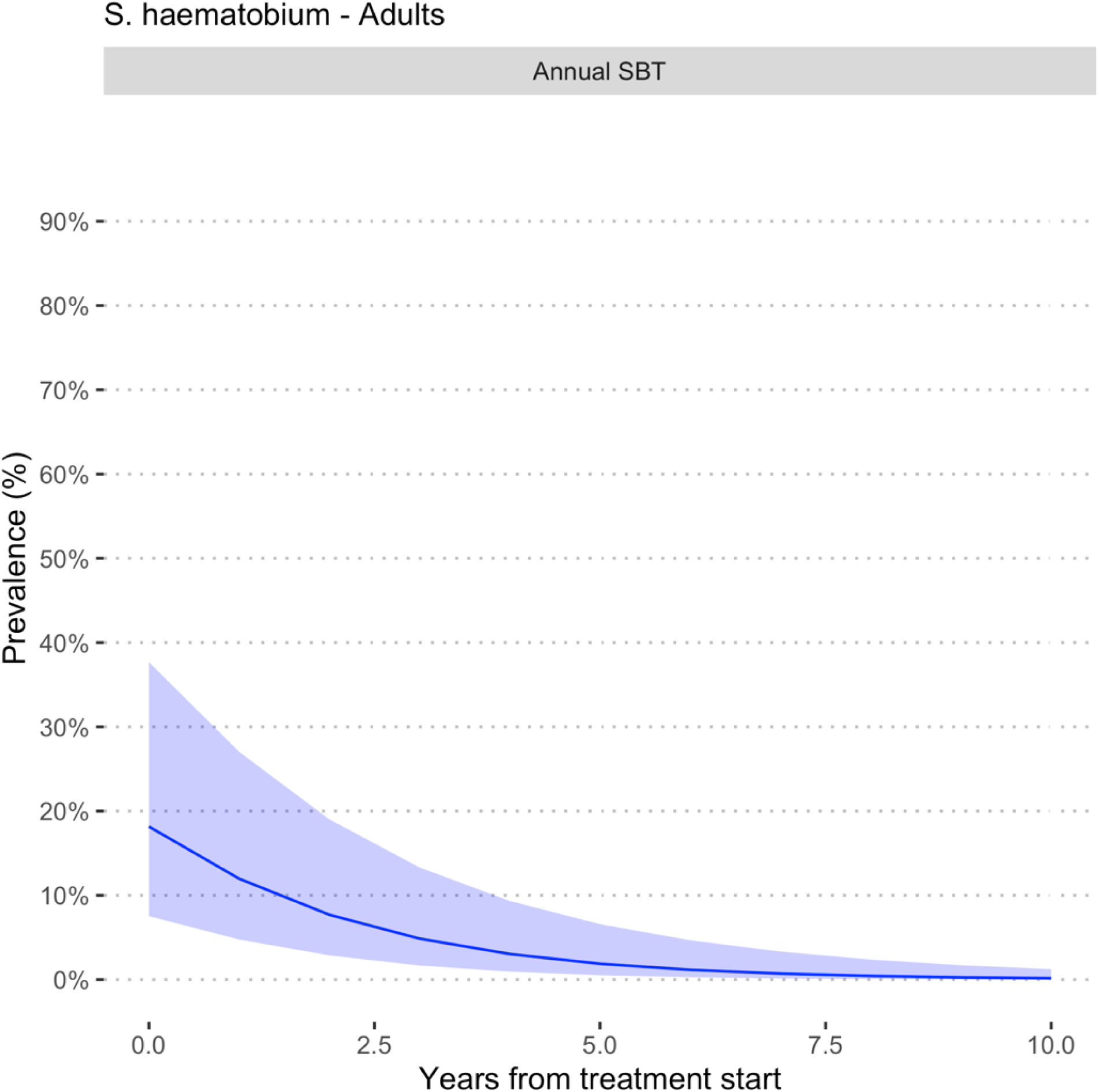
Estimated prevalence at baseline and at different follow-up times during MDA involving Adults for *S. haematobium*. All the studies used PZQ 40 mg/kg administered as a single oral dose alone or in combination with Albendazole (400 mg) in MDA campaigns in endemic countries in Sub-Saharan Africa.

**Fig. S4.**
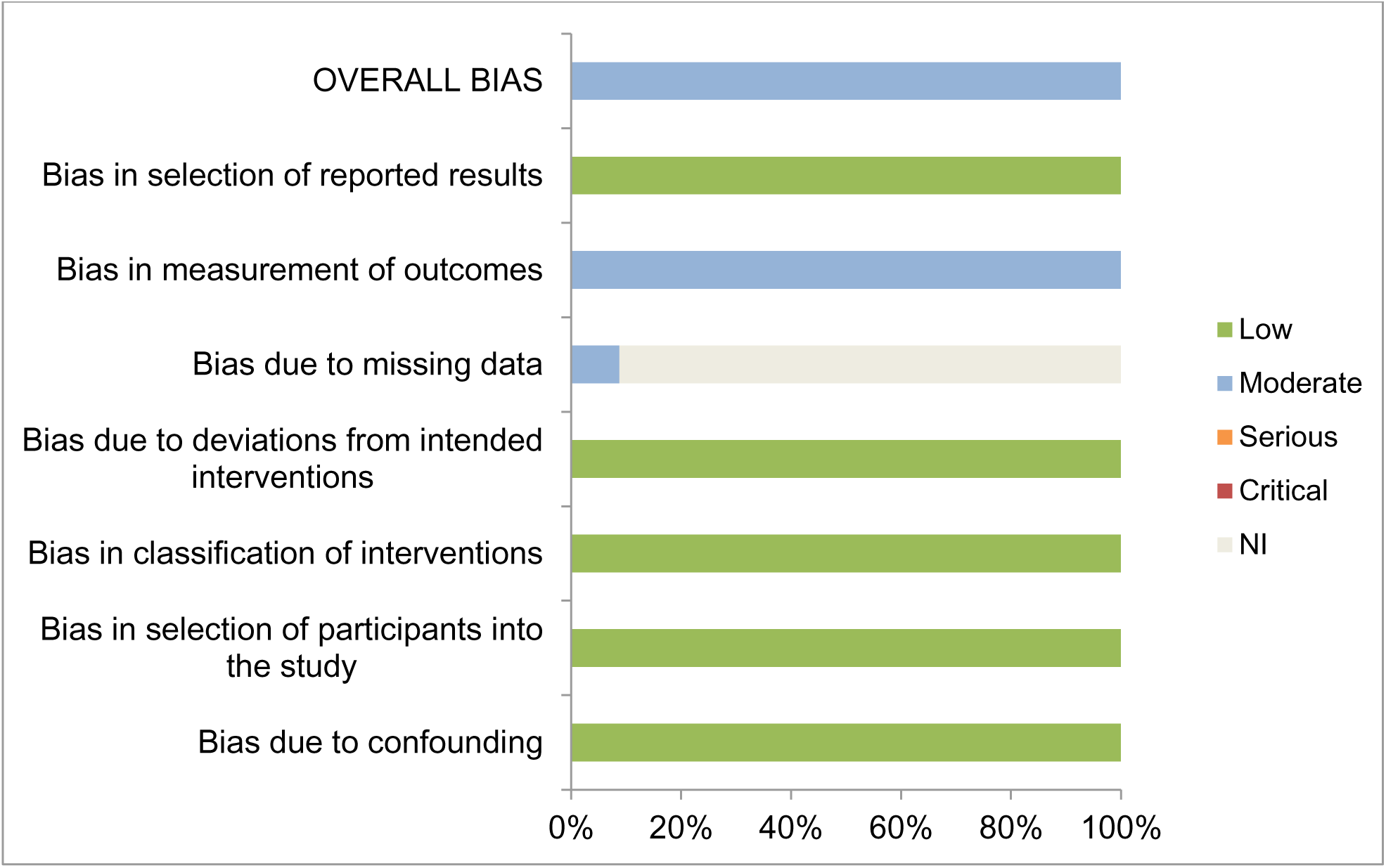
Graphical representation of Risk of bias in the included studies in the systematic review and meta-analysis on seven main domains where bias could be introduced into the study. NI= no information

**Table 4.** Results of risk of bias in assessed on seven main domains in the studies included in the systematic review and meta-analysis

| Study | Confounding | Selection of participants | Classification of interventions | Deviations from intended interventions | Missing data | Measurement of outcomes | Selection of reported results | OVERALL BIAS |
| --- | --- | --- | --- | --- | --- | --- | --- | --- |
| Abudho et al., 2018<sup>38</sup> | Low | Low | Low | Low | NI* | Moderate | Low | Moderate |
| Assare et al., 2016<sup>26</sup> | Low | Low | Low | Low | Moderate | Moderate | Low | Moderate |
| Bah et al., 2019<sup>27</sup> | Low | Low | Low | Low | NI | Moderate | Low | Moderate |
| Brinkmann et al., 1988<sup>28</sup> | Low | Low | Low | Low | NI | Moderate | Low | Moderate |
| Bronzan et al., 2015<sup>37</sup> | Low | Low | Low | Low | Moderate | Moderate | Low | Moderate |
| Hamidou et al., 2018<sup>31</sup> | Low | Low | Low | Low | NI | Moderate | Low | Moderate |
| Hodges et al., 2012<sup>34</sup> | Low | Low | Low | Low | NI | Moderate | Low | Moderate |
| Karanja et al., 2017<sup>39</sup> | Low | Low | Low | Low | NI | Moderate | Low | Moderate |
| Knopp et al., 2019<sup>46</sup> | Low | Low | Low | Low | NI | Moderate | Low | Moderate |
| Lee et al., 2015<sup>56</sup> | Low | Low | Low | Low | NI | Moderate | Low | Moderate |
| Moussa et al., 2016<sup>32</sup> | Low | Low | Low | Low | NI | Moderate | Low | Moderate |
| Mwandawiro et al., 2019<sup>40</sup> | Low | Low | Low | Low | NI | Moderate | Low | Moderate |
| Mwinzi et al., 2012<sup>41</sup> | Low | Low | Low | Low | NI | Moderate | Low | Moderate |
| Njenga et al., 2014<sup>42</sup> | Low | Low | Low | Low | NI | Moderate | Low | Moderate |
| Olsen et al., 2018<sup>47</sup> | Low | Low | Low | Low | NI | Moderate | Low | Moderate |
| Onkanga et al., 2016<sup>43</sup> | Low | Low | Low | Low | NI | Moderate | Low | Moderate |
| Ouedraogo et al., 2016<sup>24</sup> | Low | Low | Low | Low | NI | Moderate | Low | Moderate |
| Phillips et al., 2017<sup>54</sup> | Low | Low | Low | Low | Moderate | Moderate | Low | Moderate |
| SCI-supported MDA 1<sup>25</sup> | Low | Low | Low | Low | NI | Moderate | Low | Moderate |
| SCI-supported MDA 2<sup>52</sup> | Low | Low | Low | Low | NI | Moderate | Low | Moderate |
| SCI-supported MDA 3<sup>51</sup> | Low | Low | Low | Low | NI | Moderate | Low | Moderate |
| SCI-supported MDA 4<sup>36</sup> | Low | Low | Low | Low | NI | Moderate | Low | Moderate |
| SCI-supported MDA 5<sup>53</sup> | Low | Low | Low | Low | NI | Moderate | Low | Moderate |
| SCI-supported MDA 6<sup>29</sup> | Low | Low | Low | Low | NI | Moderate | Low | Moderate |
| SCI-supported MDA 7<sup>30</sup> | Low | Low | Low | Low | NI | Moderate | Low | Moderate |
| SCI-supported MDA 8<sup>55</sup> | Low | Low | Low | Low | NI | Moderate | Low | Moderate |
| SCI-supported MDA 9<sup>48</sup> | Low | Low | Low | Low | NI | Moderate | Low | Moderate |
| SCI-supported MDA 10<sup>50</sup> | Low | Low | Low | Low | NI | Moderate | Low | Moderate |
| Secor et al., 2020<sup>44</sup> | Low | Low | Low | Low | NI | Moderate | Low | Moderate |
| Senghor et al., 2016<sup>33</sup> | Low | Low | Low | Low | NI | Moderate | Low | Moderate |
| Sesay et al., 2014<sup>35</sup> | Low | Low | Low | Low | NI | Moderate | Low | Moderate |
| Shen et al., 2019<sup>57</sup> | Low | Low | Low | Low | NI | Moderate | Low | Moderate |
| Sircar et al. 2018<sup>45</sup> | Low | Low | Low | Low | NI | Moderate | Low | Moderate |
| Zhang et al., 2007<sup>49</sup> | Low | Low | Low | Low | NI | Moderate | Low | Moderate |
| **Low** | 34 | 34 | 34 | 34 | 0 | 0 | 34 | 0 |
| **Moderate** | 0 | 0 | 0 | 0 | 3 | 34 | 0 | 34 |
| **Serious** | 0 | 0 | 0 | 0 | 0 | 0 | 0 | 0 |
| **Critical** | 0 | 0 | 0 | 0 | 0 | 0 | 0 | 0 |
| **\* No information (NI)** | 0 | 0 | 0 | 0 | 31 | 0 | 0 | 0 |

